# Characteristics of dynamic assessments of word reading skills and their implications for validity: A systematic review and meta-analysis

**DOI:** 10.1101/2023.03.20.23287486

**Authors:** Emily Wood, Kereisha Biggs, Monika Molnar

## Abstract

Dynamic assessments (DAs) of word reading skills demonstrate strong criterion reference validity with word reading measures (WRMs). However, DAs vary in the skills they assess, their format and administration method, and the type of words and symbols used in test items. These characteristics may have implications on assessment validity. To compare validity of DAs of word reading skills on these factors of interest, a systematic search of five databases and the grey literature was conducted. We identified 35 studies that met the inclusion criteria of evaluating participants aged 4-10, using a DA of word reading skills and reporting a Pearson’s correlation coefficient as an effect size. A random effects meta-analysis with robust variance estimation and subgroup analyses by DA characteristics was conducted. There were no significant differences in mean effect size based on administration method (computer vs. in-person) or symbol type (familiar vs. novel). However, DAs that evaluate phonological awareness or decoding (vs. sound-symbol knowledge), those that use a graduated prompt format (vs. test-teach-retest), and DAs that use nonwords (vs. real words) demonstrated significantly stronger correlations with WRMs. These results inform selection of DAs in clinical and research settings, and development of novel, valid DAs of word reading skills.

## Introduction

Literacy is a complex construct requiring integration of multiple skills. Simply, it can be described as the product of the ability to recognize or decode words and to comprehend language (Hoover & Gough, 1990). In this review, we derive our definition of the construct of word reading skills from the subskills that comprise word recognition ability in the evidence-based model Scarborough’s reading rope (Scarborough, 2001). These subskills – phonological awareness, knowledge of the alphabetic principle (or sound-symbol knowledge) decoding and sight word recognition have been consistently found to be among the strongest and most accurate predictors of reading ability for young children (e.g., Catts et al., 2005; Ehri, 1998; Hogan et al., 2005). In speech-language pathology (SLP), psychology and education, most word reading tools employ a static assessment (SA) paradigm (e.g., Phonological Awareness Test-2 (PAT-2:NU), Robertson & Salter, 2017; Woodcock Reading Mastery Test-III(WRMT-III), Woodcock, 2011). In SA, an examiner passively evaluates performance without providing prompts or feedback to measure a child’s acquired knowledge in a given domain (Grigorenko & Sternberg, 1998). Children from, diverse linguistic backgrounds or those with fewer literacy experiences are prone to perform poorly because they have limited or different acquired knowledge compared to the English monolingual children for whom the tests are designed (Bedore & Peña, 2008; Ginsborg, 2006). When many children underperform, it leads to floor effects, rendering it difficult to discern those who are truly at-risk from those who have had insufficient linguistic or educational experiences. This can result in failure to identify word reading difficulties early (Catts et al., 2009).

Given these limitations, interest in alternative approaches, like dynamic assessments (DAs), has been increasing. While SAs measure a child’s acquired skills DAs examine a child’s ability to learn a skill with support in the form of teaching, feedback and prompting from the examiner in the test (Grigorenko & Sternberg, 1998). This approach reduces bias and misidentification of difficulty because the impact of previous linguistic or literacy experiences on test outcomes is minimized (Bedore & Peña, 2008; Petersen & Gillam, 2013). Reviews that have evaluated the use of DAs report promising findings on their utility and validity. DAs demonstrate greater predictive validity than SAs across several domains (e.g., DAs of cognitive ability, literacy, and mathematics) (Caffrey et al., 2008). DAs of word reading can predict unique variance in later reading ability beyond SAs (Dixon et al., 2022a), can contribute to the accurate identification of reading difficulties (Dixon et al., 2022b), and demonstrate consistent validity with word reading outcome measures across typically developing, at-risk, bilingual, and monolingual children (Wood et al., 2023). However, these DAs are also characterized by heterogeneity in terms of the word reading skills they assess, their format, administration method; in addition to the word and symbol type they use.

Previous reviews have explored the impact of these factors in the domain of static assessment. For example, the factor of administration method (virtual vs. in-person) was considered by Alfano et al., (2022). Authors found that across the 7 included studies, there were no significant differences between online and in-person administration of pediatric language and literacy assessments. Outcomes such as these provide support for the development of novel SA tools that can be administered online. Whether administration method, or any other of the characteristics described below affect the validity of DAs has not been considered thus far. In this meta-analysis we will directly examine characteristics of DAs and determine which if any types of assessments are superior to others in terms of their criterion reference validity with word reading measures. Outcomes have implications for the revision of existing assessments and development of novel DAs. The characteristics of interest are described in more detail below.

### Word Reading Skill Type

DAs have been developed to evaluate various skills associated with literacy development and ability including decoding (e.g., Cho et al., 2017), phonological awareness (e.g., Gellert & Elbro, 2017b), sound-symbol knowledge (e.g., Clayton et al., 2018), morphological awareness (e.g., Navarro et al., 2018), expressive vocabulary development (Peña et al., 2001), oral narratives (Peña et al., 2014), reading comprehension (e.g., Gruhn et al., 2020) and working memory (e.g., Swanson, 1994). As stated, in this review, we focus exclusively on DAs of that evaluate the word reading skills of decoding, phonological awareness or sound-symbol knowledge because these skills have consistently been found to be among the strongest predictors of word reading ability (e.g., The National Literacy Panel (2008)).

Historically research in the validity of static measures of these word reading skills has found that all three correlate strongly with later word reading ability. The National Early Literacy Panel (2008) which included nearly 300 studies, and a meta-analysis of 60 effect sizes by Elbro and Scarborough (2003) both documented strong correlations between kindergarten letter(-sound) knowledge (*r*=.50 and *r*=.52 respectively) and decoding (*r*=.53 and *r*=.57 respectively) and later word reading ability. Similarly, the National Early Literacy Panel and a meta-analysis of 235 studies by Melby-Lervåg et al., (2012) both found moderate-strong correlations between kindergarten-aged phonemic awareness ability, a subskill of phonological awareness and later reading ability, (*r*=.42 and *r*=.43 respectively). These across-study results suggest that SAs of word reading skills are all similarly correlated with later reading abilities, with slight advantage to decoding and letter-sound knowledge over phonological awareness. Whether this is indeed the case in DA is investigated by the current study.

In the realm of DA there have been fewer systematic examinations into the capacity of these skills to predict later word reading. A recent systematic review of 18 studies found that across studies, DAs of phonological awareness and decoding predicted between 1-21% additional unique variance beyond traditional static measures in later word reading ability, but that a DA of paired associate learning (a task akin to learning sound-symbol knowledge) accounted for only 6% unique variance (Dixon et al., 2022a), suggesting a different pattern of prediction between DAs and SAs. Given that DAs evaluate ability to learn rather than acquired knowledge, it may be that assessments that evaluate more complex skills better permit children to demonstrate their ability to learn in the context of DA. Simple sound-symbol knowledge tasks require that a child learn the name that corresponds to a symbol.

However, complex phonological awareness tasks like phoneme substitution require that a child identify a sound, delete it, replace it with a new sound and blend the new sounds together to form the new word, and complex decoding tasks require integration of both sound-symbol knowledge and phonological awareness skills. These more complex tasks might be better suited to capturing learning potential in the context of DA. Given this, we expect that DAs of decoding and phonological awareness may demonstrate stronger correlational relationships with WRMs, relative to DAs of SSK.

### Format

DAs come in many formats, but there are two primary approaches (Lantolf & Poehner, 2004). Interactionist DA is unscripted and endeavors to modify cognitive or skill ability. The examiner responds contingently to the individual examinee and their capacities. Interventionist DA, however, more closely parallels SA. The examiner provides pre-defined, non-contingent and increasingly explicit levels of support in response to student performance. Its scripted nature requires less clinical skill and time to administer, and its standardization permits evaluation of its psychometric validity (Poehner, 2008). Interventionist DAs across cognitive domains demonstrate stronger predictive validity than interactionist DAs (Caffrey et al., 2008). In the field of word reading assessment, all DAs can be characterized as interventionist. The studies included in this review focus on two common formats of interventionist DA.

The first, referred to as the (test)/teach/retest (TT) format in this paper, consists of a static pre-test, followed by a dynamic teaching phase, and a static re-test (Budoff, 1987). During the teaching phase, children receive feedback and instruction. Not all assessments incorporate the initial static pre-test. If one is conducted, post-test performance is compared to the initial score to assess the difference in performance following teaching. When no pre-test is administered, the post-test measures how a child performs after receiving explicit dynamic instruction in a task.

The second approach, referred to as the graduated prompts (GP) format in this paper, combines teaching and testing phases of the assessment within each item (Brown & Ferrara, 1985). Children are provided with feedback regarding their response. If incorrect, a hierarchy or series of increasingly explicit prompts are provided, until the child answers correctly, or all prompts are exhausted. The greater number of prompts required, the lower the score on an item (Brown & Ferrara, 1985). A previous review suggested that there are no differences in classification accuracy of reading disorder for DAs that use a GP vs. TT format and that both formats are used with similar frequency in assessment of word reading (Dixon et al., 2022b).

Given that previous research has found that noncontingent DAs correlate more strongly with word reading outcomes than contingent ones (Caffrey et al., 2008), we anticipate that there DAs that use a GP format would be superior to those that use the TT format, because this approach is highly explicit and structured, while TT allows greater flexibility in response to the student in the training/teaching portion. However, to date no quantitative comparison of the validity of these two interactionist formats has been conducted.

### Administration Method

Assessments, dynamic or otherwise, can be conducted in-person or via computer. Development of virtual or computer-based assessments has become increasingly important, in the wake of the COVID-19 pandemic and the subsequent shift to distance learning (Campbell & Goldstein, 2022). No significant differences have been found between administering an SA online vs. in-person (Alfano et al., 2022), but this factor has not been considered in the context of DAs, which can be administered in-person (e.g., Spector, 1992), virtually by an examiner (e.g., Barker & Saunders, 2020), or in a computer program where no examiner is required (e.g., Aravena et al., 2018). A recent review found that computerised DAs were used less frequently than in-person measures (Dixon et al., 2022b), but the implications of administration method on the validity of DAs have not yet been considered quantitatively. It is anticipated that because DA is characterized by increased interaction between examiner and examinee, it maybe be impacted to a greater extent by computer administration than static assessment. Importantly, post-pandemic, many clinicians and researchers continue to operate virtually, and therefore the factor of administration method and its implications on validity should be considered.

### Word Type

Assessments of the word reading skills of phonological awareness and decoding use either real words or nonwords, which are made-up words that abide by the language’s phonotactic and orthotactic constraints (e.g., “meeb” in English). Commercially developed SA tools such as the Comprehensive Test of Phonological Processing –2 (CTOPP-2, Wagner et al., 2013) which evaluates phonological awareness, or the Woodcock Reading Mastery Test – Third Edition (WRMT-III, Woodcock, 2011), which evaluates decoding, include subtests with words and nonwords. Some DAs like the CUBED-3 dynamic test of decoding include a word reading and nonword decoding measure (Petersen et al., 2016). However, many DAs have fewer subtests and tend to employ either words or nonwords. For example, Gellert and Elbro’s dynamic phoneme identification task uses real words (2017b), but their dynamic decoding measure uses nonwords (2017a).

Reading words and non-words are purported to tap into different processes (Shapiro et al., 2013). Children may initially recognize familiar words by sight without activating their knowledge of sound-symbol correspondences, phoneme blending and decoding skills (Ehri & Wilce, 1985). For example, recognizing their name or a high frequency word like “the” in print. However, when reading nonwords, decoding skills are necessary because these words are unfamiliar (Hoover & Tunmer, 1993). Similarly, word and nonword phonological awareness tasks may activate different skills. When using real words, performance may be impacted by acquired vocabulary knowledge, while nonword tasks may be a purer form of phonological ability (Wagner et al., 2013).

In the domain of oral language assessment, nonword repetition tasks have been shown to reduce bias against culturally and linguistically diverse children (Ortiz, 2021). Nonword decoding and phonological awareness tasks may similarly reduce bias against those with different or limited literacy experiences in word reading skills assessments. Children enter kindergarten with a wide range of language and literacy abilities that can be attributed linguistic diversity, their home literacy environment, access to books and libraries, or exposure to literacy instruction in preschool (Ackerman & Barnett, 2005). Importantly, nonword tasks do not disadvantage strong readers with advanced lexical knowledge (Castles et al., 2018). They can account for significant unique variance in word reading ability beyond real word reading (e.g., Hogan et al., 2005). To date, no studies have considered the role of word type in validity of DAs of word reading skills. We expect that nonword tasks will be better suited to predict later reading ability in a DA paradigm, because DAs are designed to measure ability to learn a skill. Because nonword items are unfamiliar, they require learning to complete the word reading skill task.

### Symbol Type

Word reading assessments of sound-symbol knowledge (SSK) and decoding use either familiar or novel symbols. Typically, SAs use the letters or characters of the language for which they were created. For instance, in the PAT-2 (Robertson & Salter, 2017) the phoneme-grapheme subtest (a measure of sound-symbol knowledge) evaluates a child’s acquired knowledge of the relationship between familiar English letters and sounds, and the phoneme decoding subtest evaluates ability to read nonwords comprised of English graphemes. Recently, there has been increased interest in using novel symbols in DAs as it permits evaluation of how well a child can learn new symbol-sound relationships (e.g., that the symbol ◊= sound /m/, Gellert & Elbro, 2017a), and apply this knowledge to decode symbol-based words (e.g., that the symbols ◊ = the nonword /ma/, Gellert & Elbro, 2017a), while minimizing the influence of previous linguistic and literacy exposure.

No prior reviews have examined whether symbol type (novel vs. familiar) affects DA’s strength of correlational relationship with word reading ability. Primary studies suggest that DAs that use novel symbols can differentiate between typical readers and those with dyslexia (Aravena et al., 2013, 2018). These measures can explain unique variance in later reading ability beyond traditional measures for preliterate children (Horbach et al., 2015). When administered in kindergarten to predict ability in grade 1, a DA decoding measure that used novel symbols (Gellert & Elbro, 2017a) had a superior diagnostic accuracy to a one that used familiar letters (Petersen et al., 2016). Use of novel symbols is a recent development in the field of word reading assessment and there has not yet been a systematic quantitative examination of the relative validity of these two approaches. We hypothesize that in a DA paradigm, evaluating ability to learn SSK or decoding skills with novel symbols may be associated with stronger criterion referenced validity again because it may permit greater capacity to evaluate ability to learn, given the novelty of the symbols used in the tasks.

### The Current Study

The current study investigates whether these characteristics of word reading skill, format, administration method, word, and symbol type affect DA’s validity as measure by association with performance on word reading measures. We examine criterion validity, as represented by the correlation between performance on a DA of word reading skills (phonological awareness, sound-symbol knowledge or decoding) and a word reading measure (single real or nonword accuracy or fluency). Like Caffrey et al., (2008) we will use Pearson’s correlation coefficients as our effect size, given that these are the most observed type of effect size reported across studies. We focus exclusively on DAs of word reading skills as they are best suited to evaluate and predict reading ability in our target demographic of children are learning to read (Catts et al., 2005). We will examine overall validity and stratify DAs into subgroups by their format (graduated prompts vs. train/test), administration method (in-person vs. via computer), word type (real word vs. nonword) and symbol type (familiar vs. novel). We also conduct a comprehensive search of the gray literature and include studies published in French and Spanish.

Outcomes of this review will inform which characteristics of DAs of word reading skills are associated with the strongest correlations with word reading measures. For clinicians, outcomes will provide insight into which DA measures are appropriate for use in their practice. For researchers, a quantitative examination how these factors affect validity of DAs of word reading skills can inform revisions of existing measures or development of new tools. This can be achieved by modifying or developing tests with characteristics shown to be most strongly associated with performance on word reading measures.

### Research Question

Do the following variables have implications on the validity of dynamic assessments of word reading skills?

A. *Word reading skill type:* Phonological awareness (PA), sound-symbol knowledge (SSK) vs. Decoding
B. *Format*: (Test)-teach-retest (TT) vs. graduated prompts (GP)
C. *Administration method*: Computer vs. in-person
D. *Word type*: Real word vs. nonwords
E. *Symbol type*: Novel vs. Familiar

## Method

The review objectives and meta-analytic approach were planned a priori and detailed in a registered protocol on the Open Science Framework (Wood & Molnar, 2022).

### Eligibility Criteria

Study inclusion criteria were determined a priori and outlined in the review protocol (Wood & Molnar, 2022). Included studies are:

i. Primary research articles found in peer-reviewed journals, or unpublished grey literature such as Masters or Doctoral theses found in preprint repositories and on Google Scholar.
ii. Studies that assessed children with a mean age between 4;0 and 10;0 who were monolingual or bi/multilingual, typically developing, at-risk for reading or diagnosed with a reading difficulty. Articles that included adults or children with other developmental challenges, such as hearing impairment, developmental language disorder, or autism spectrum disorder were excluded.
iii. Articles that reported a correlation coefficient between a DA of a word reading skill, and a word reading measure, concurrently or longitudinally.
iv. No limitation was placed on setting or location, but only articles written in English, French, Spanish, or a different language with full text translation to one of these languages were included.

### Search Strategy and Information Sources

An initial search was carried out in 5 databases, MEDLINE, Embase, CINAHL (Cumulative Index to Nursing and Allied Health Literature), PsycINFO and ERIC (Education Resources Information Centre), using the terms “dynamic assessment” and “literacy” as well as their related keywords in titles and abstracts. The search strategy was developed via consultation with a University of Toronto librarian. A complete list of search terms used in each database can be found in Tables 1 and 2 of the supplemental files. No filters were used. Equivalent terms “dynamic assessment” and “literacy” were searched in MedArxiv, EdArxiv and PsyArxiv. Forward searching was then completed on Google Scholar using the “cited by” function with included articles. To check whether any relevant articles were missed during the database, preprint, and Google Scholar search, the reference lists of the included articles were reviewed and compared to the list of included articles. Finally, appeals for unpublished work were made via social media callouts, posts to mailing lists, and direct emails to labs across Canada, the United States and Europe that reported conducting research in field of literacy.

**Table 1.**
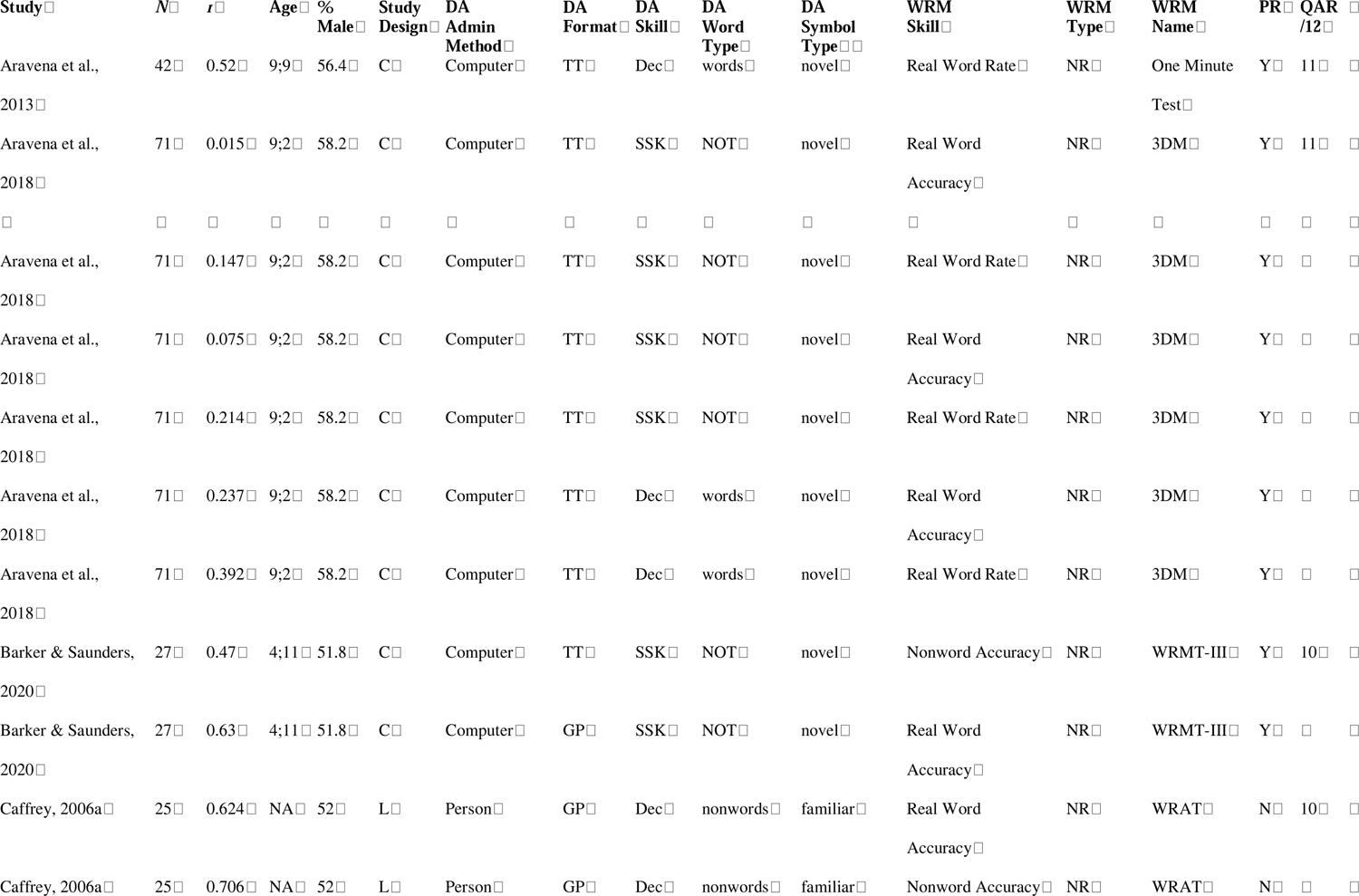
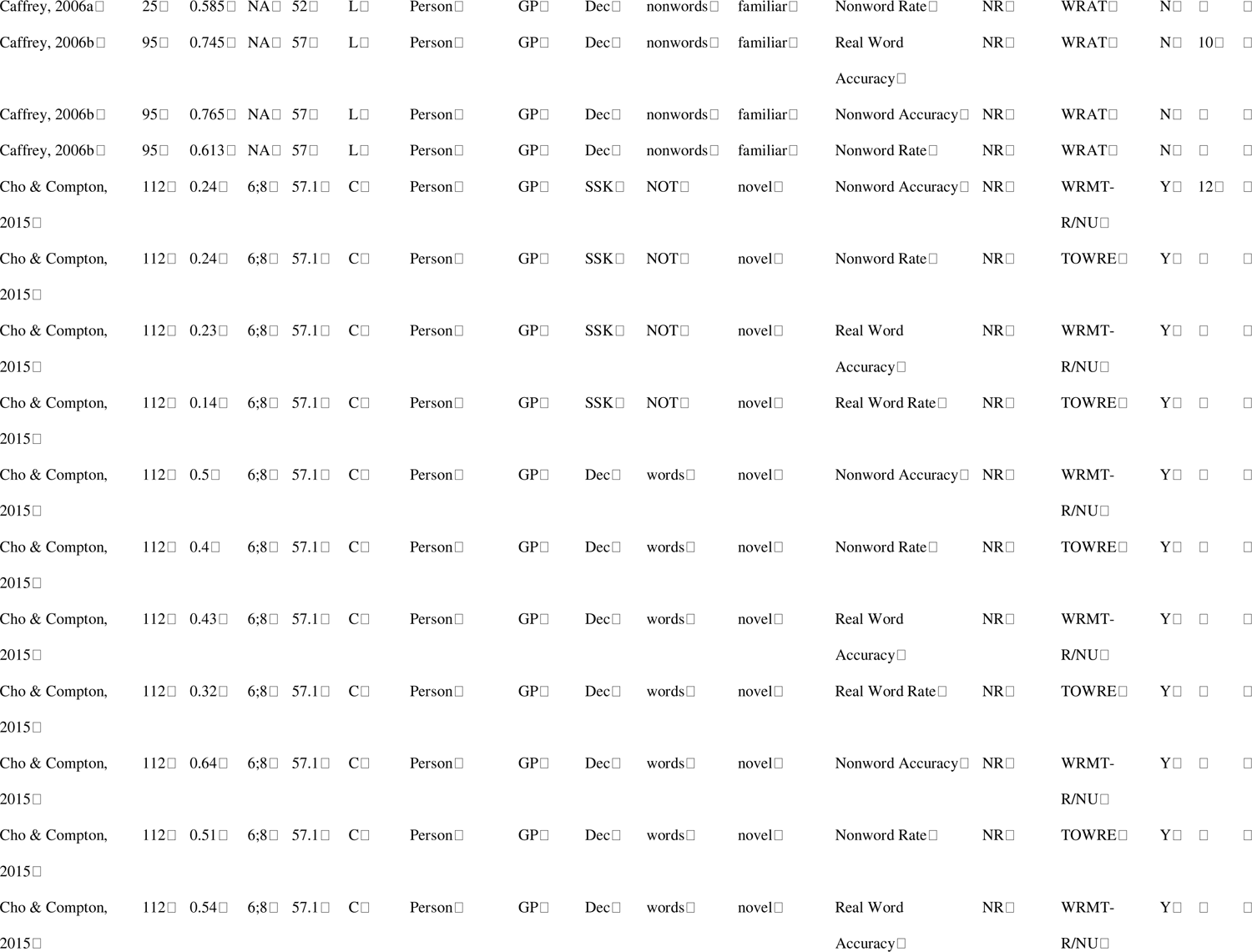
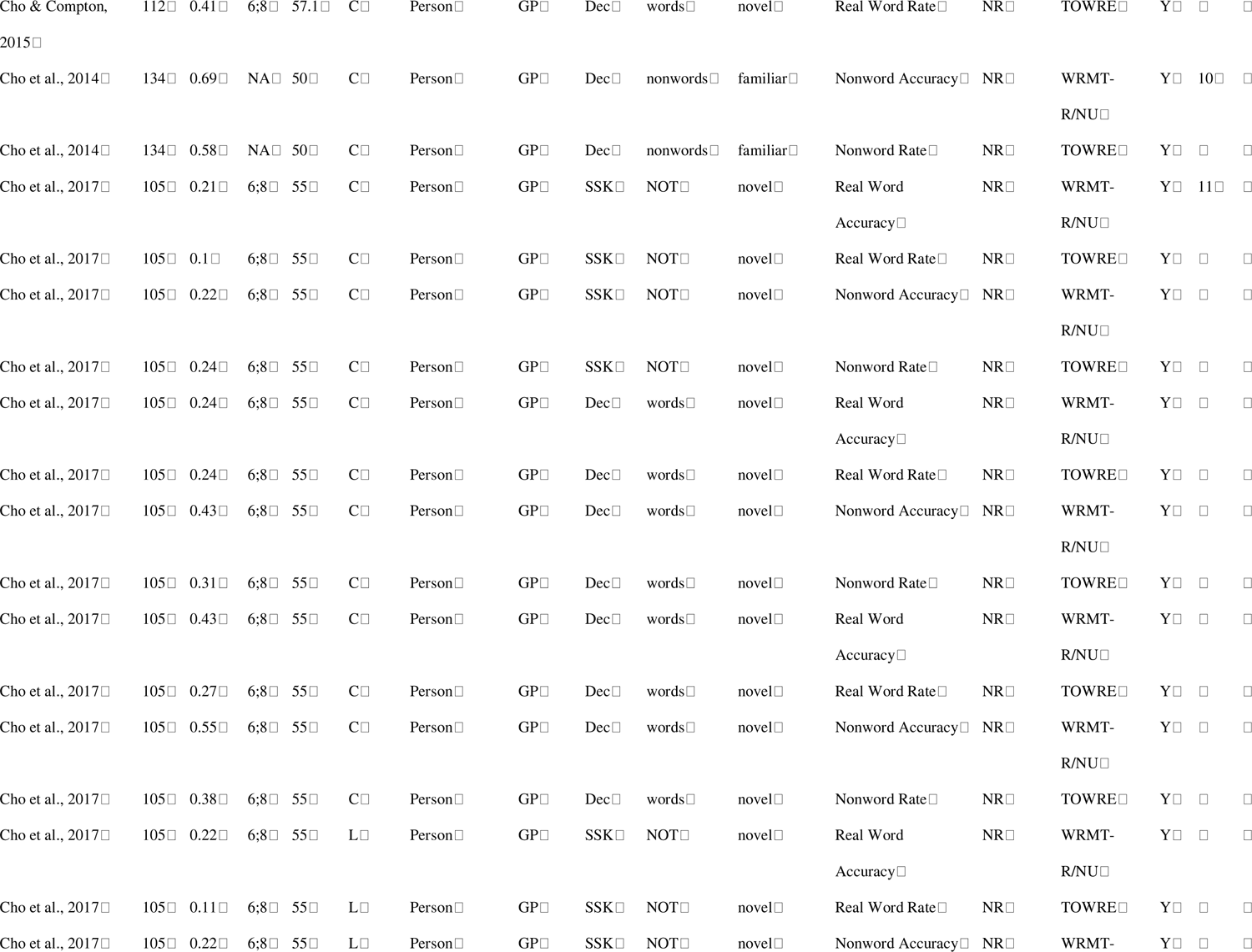
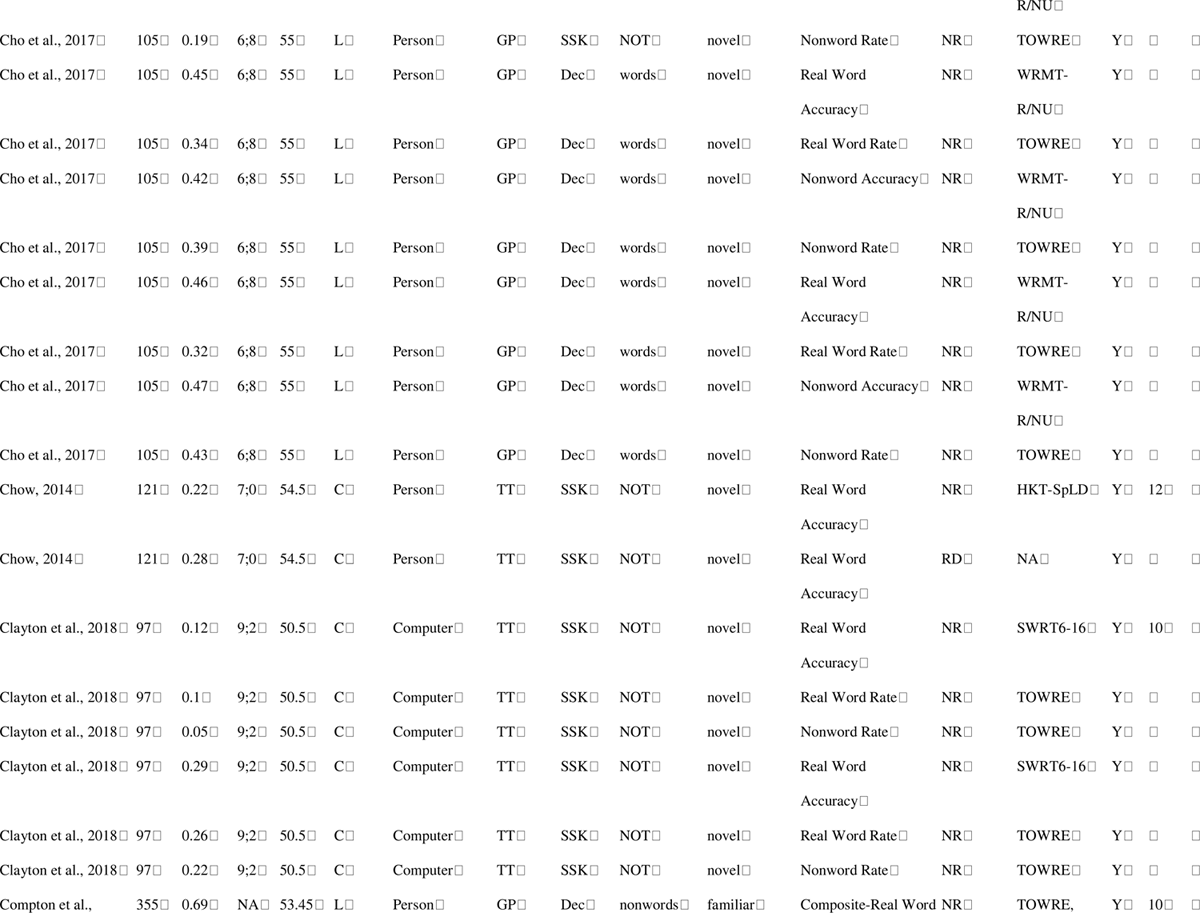
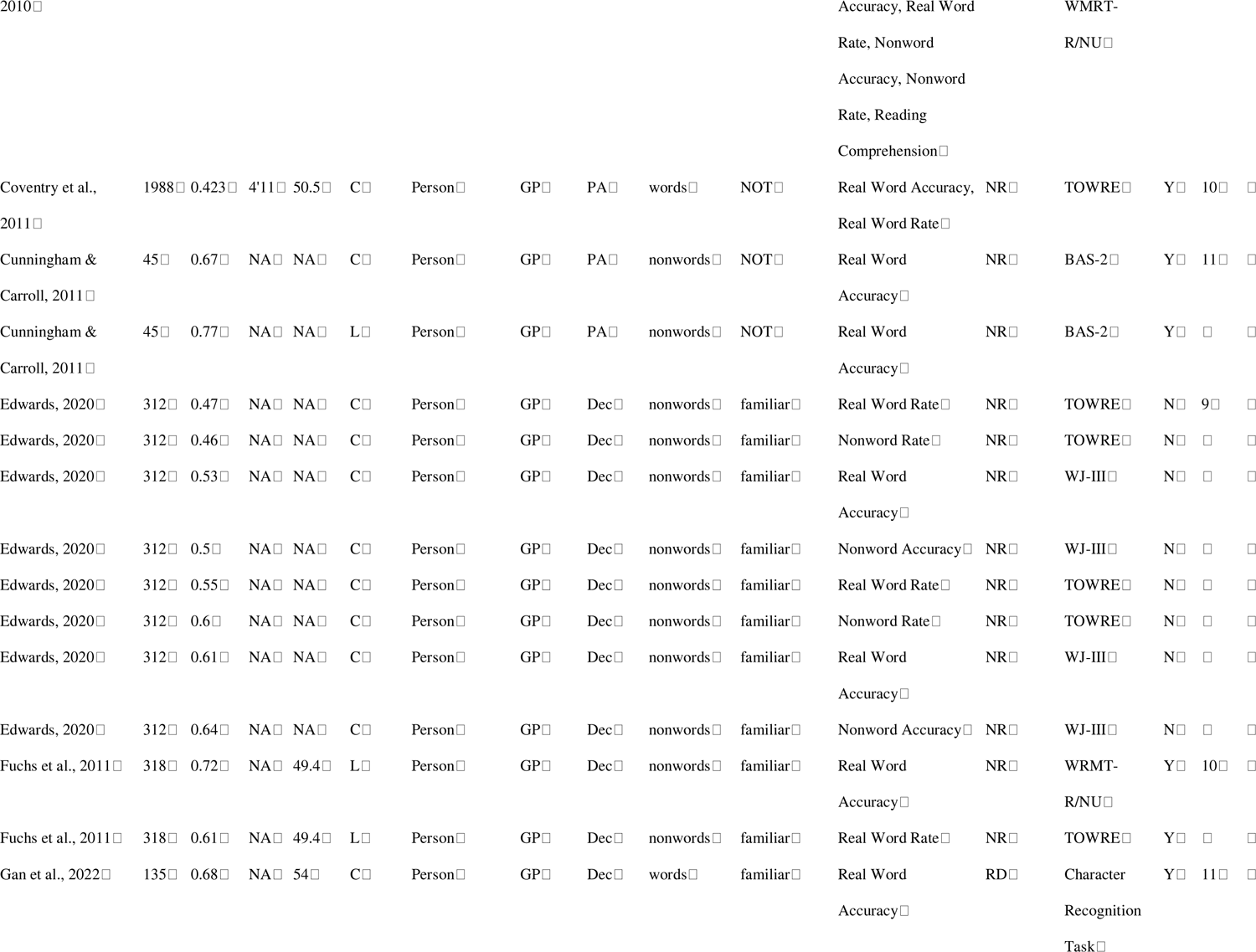
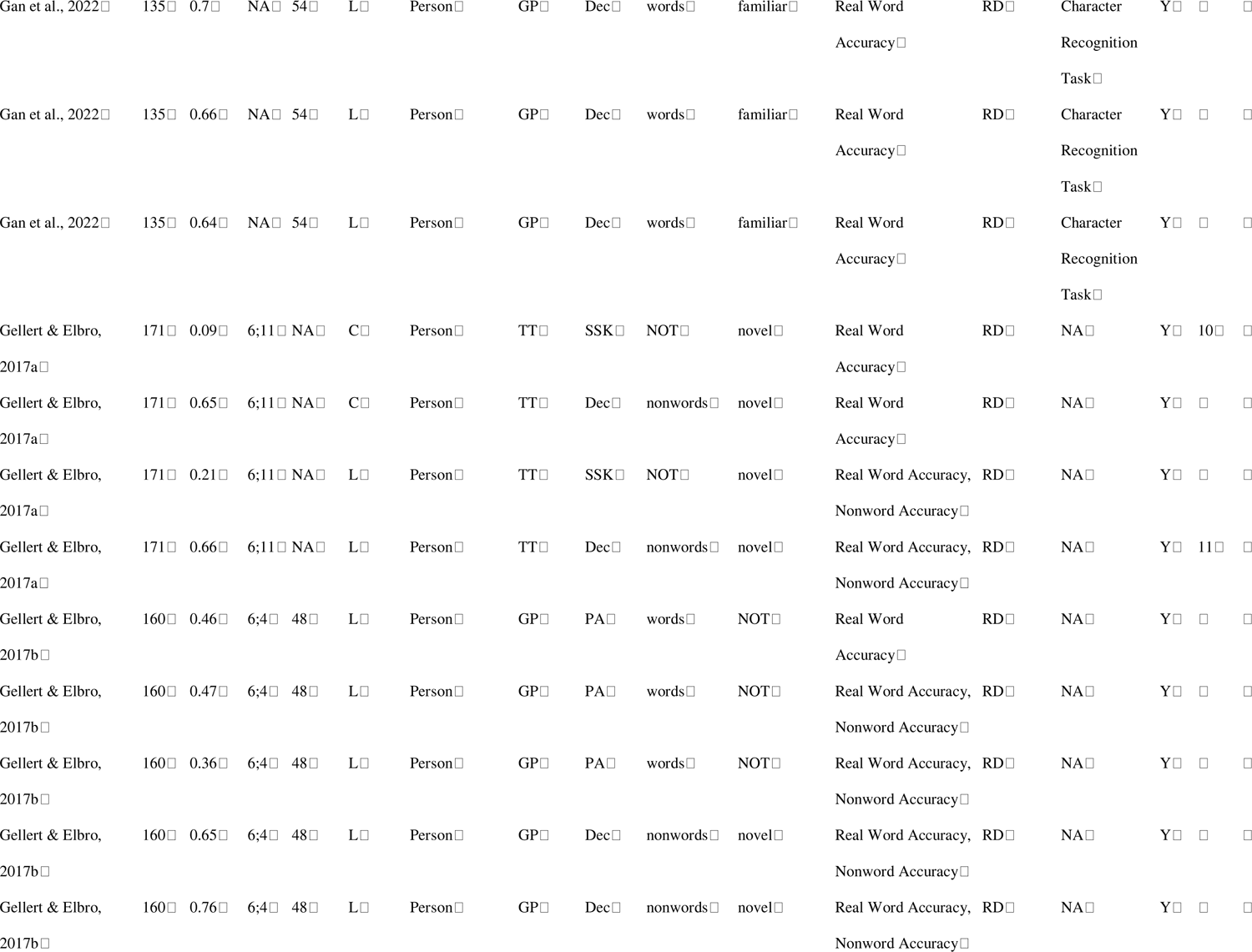
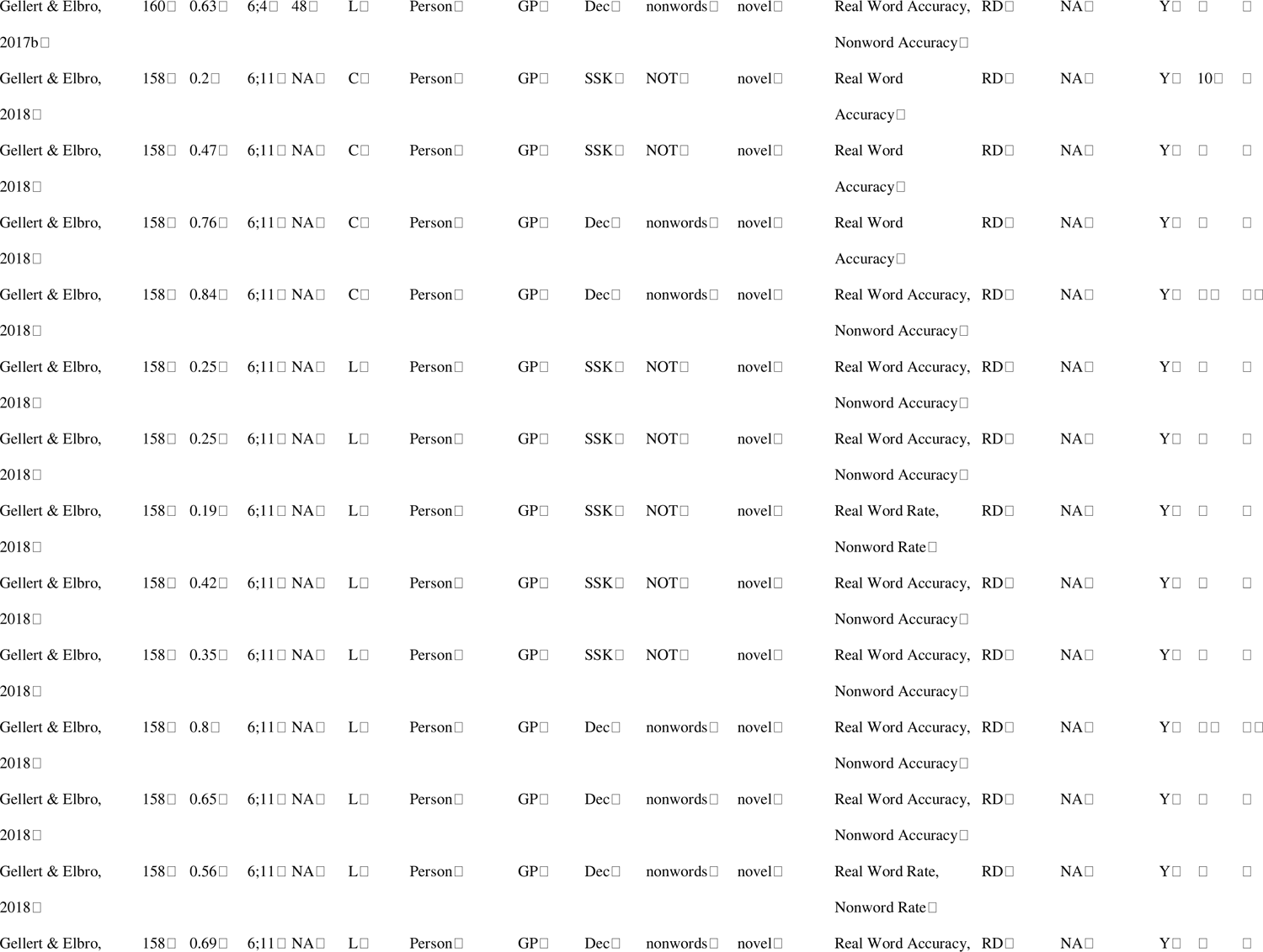
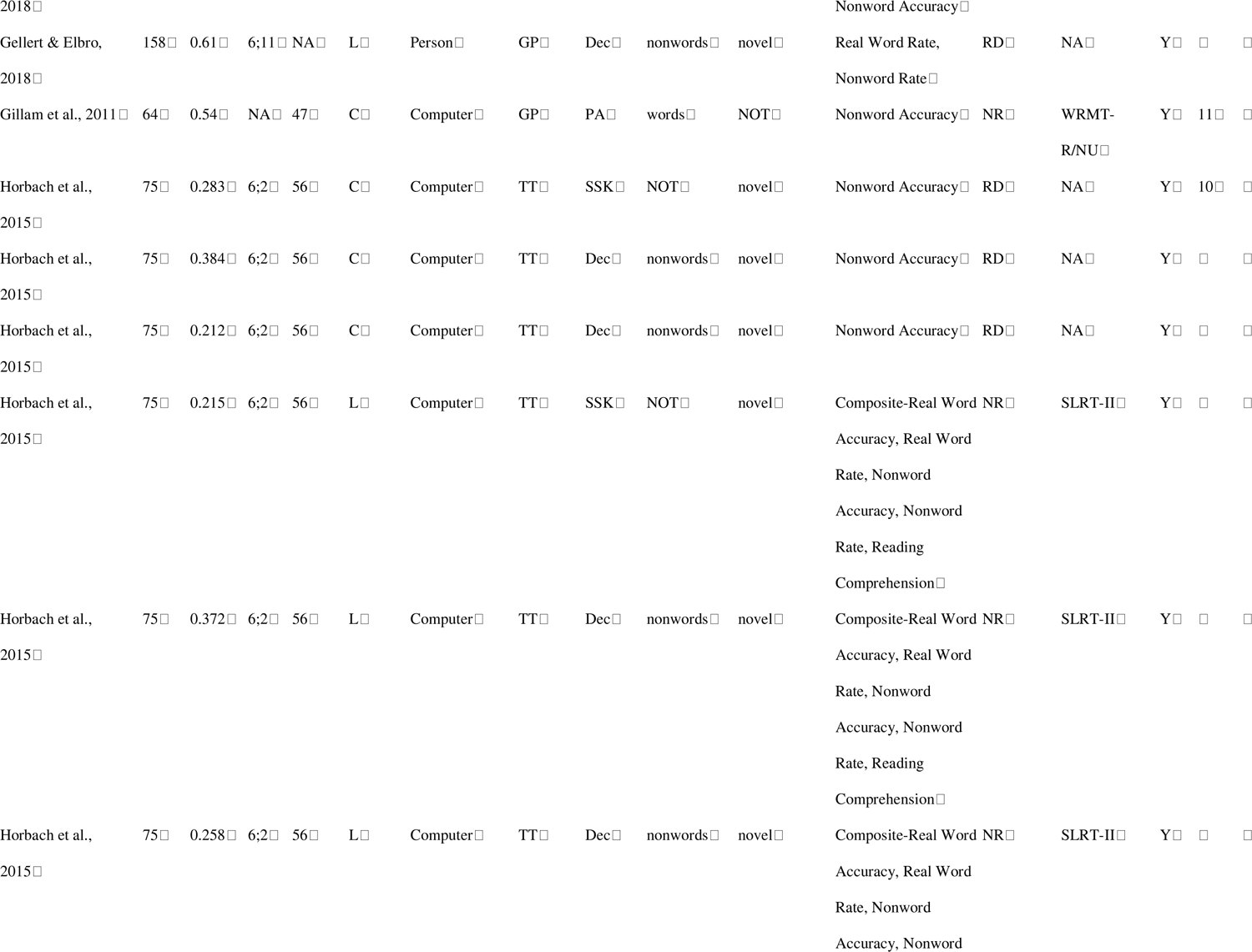
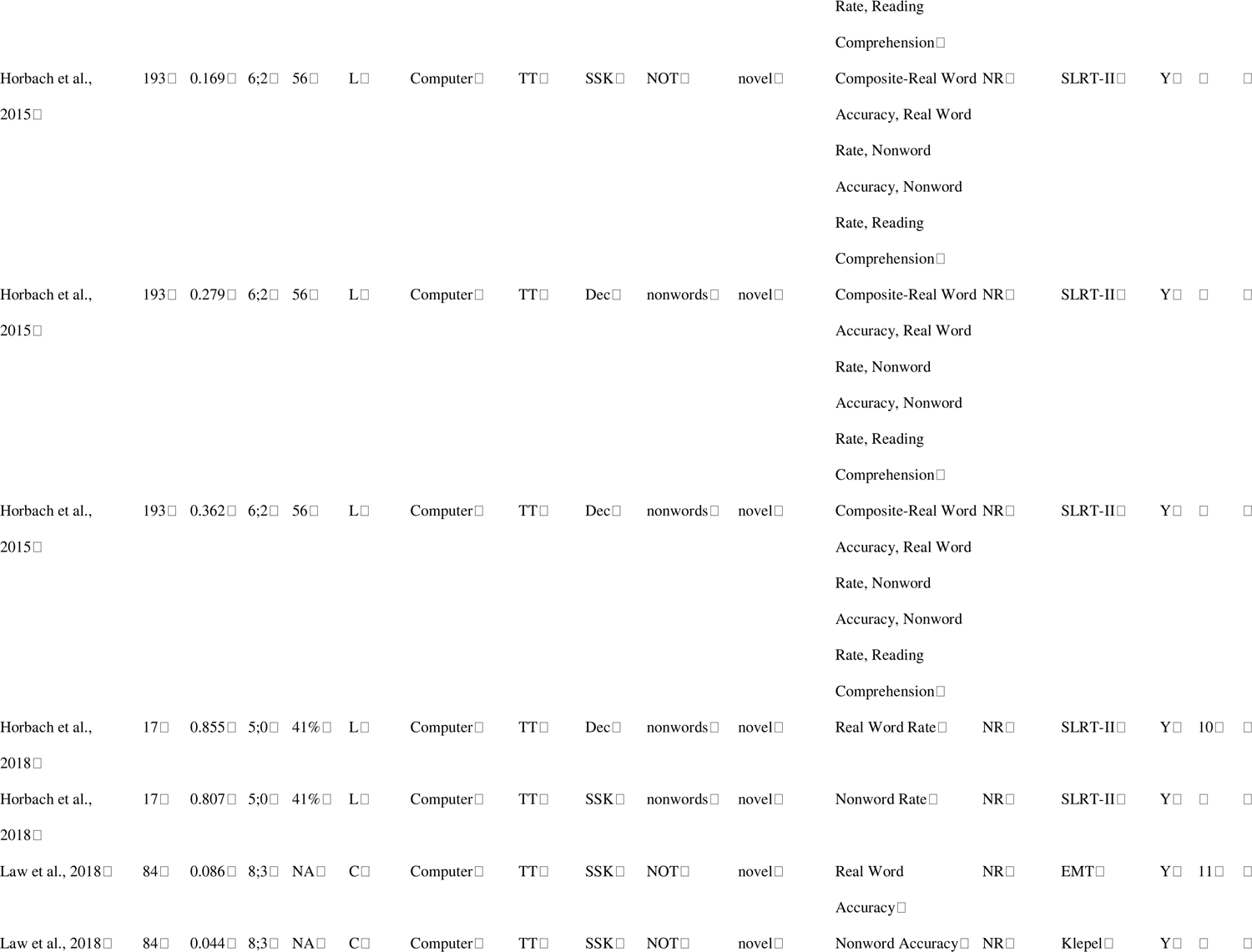
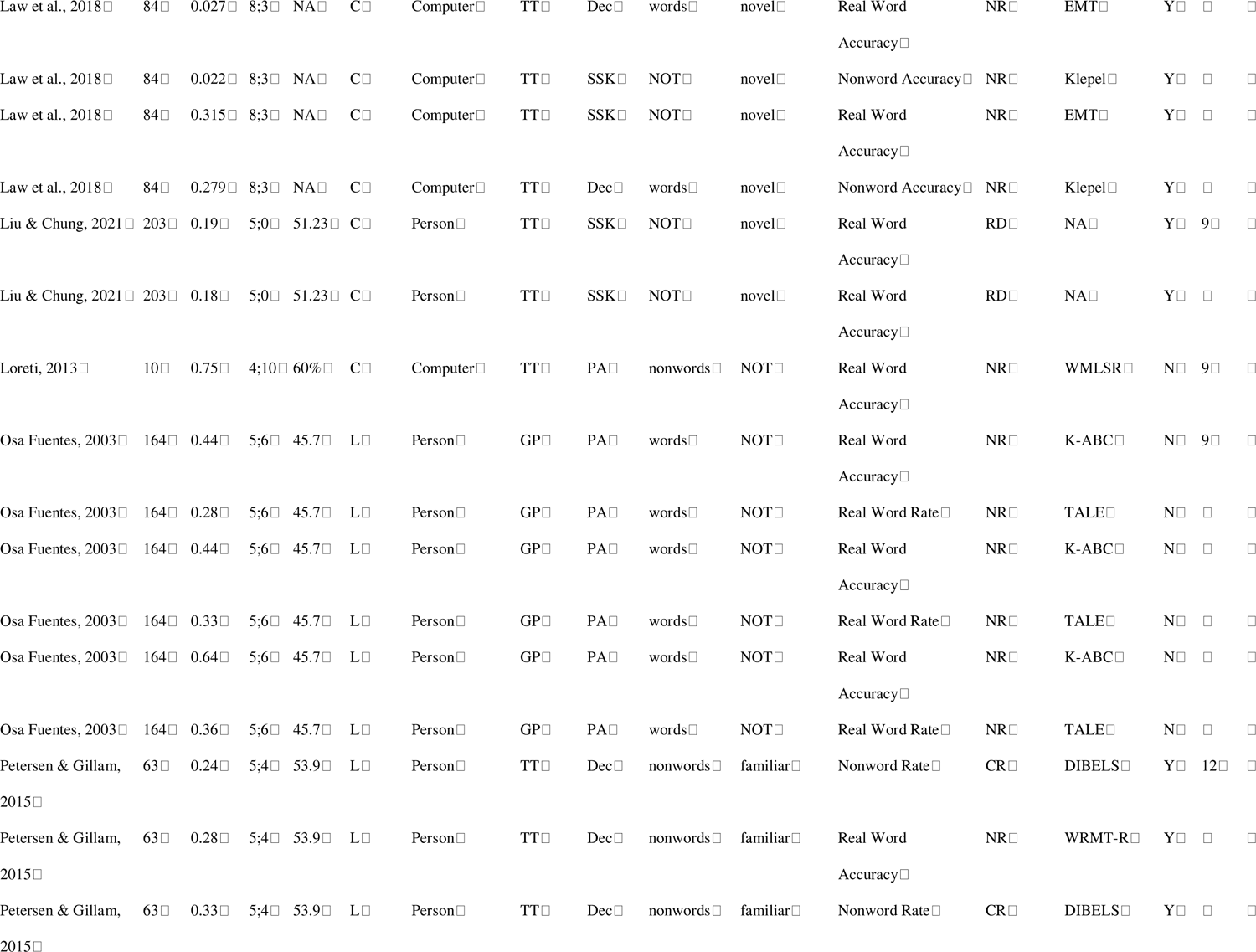
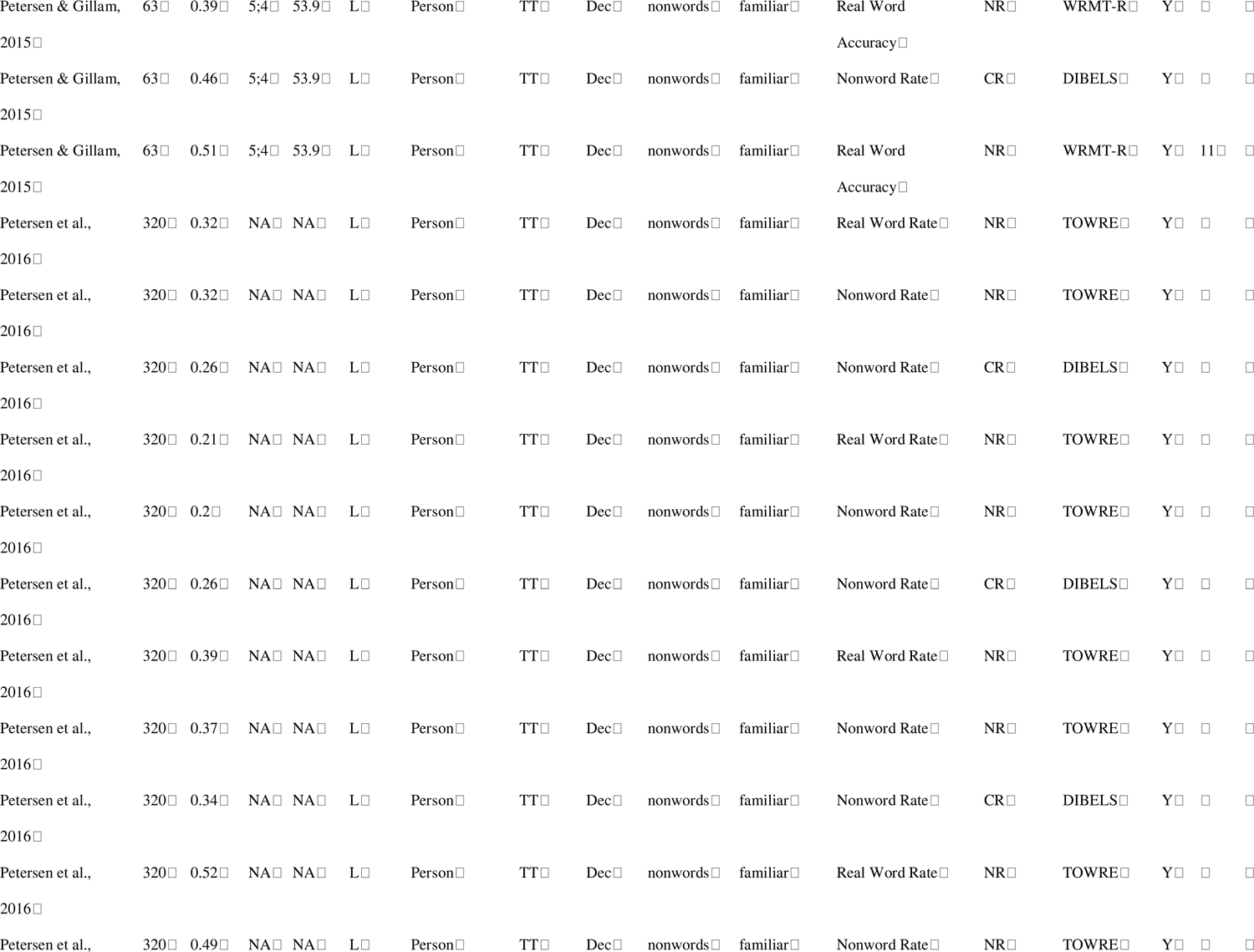
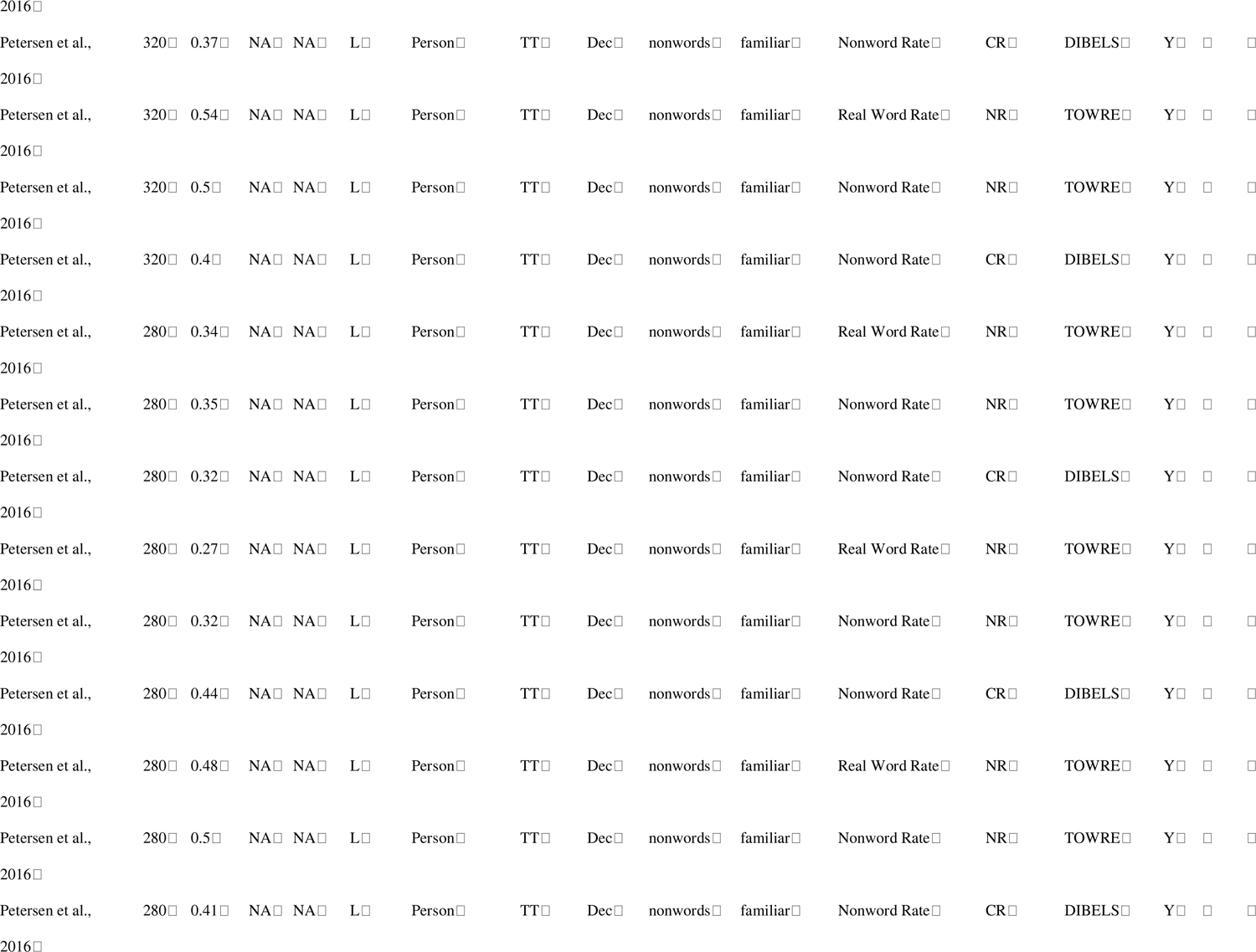
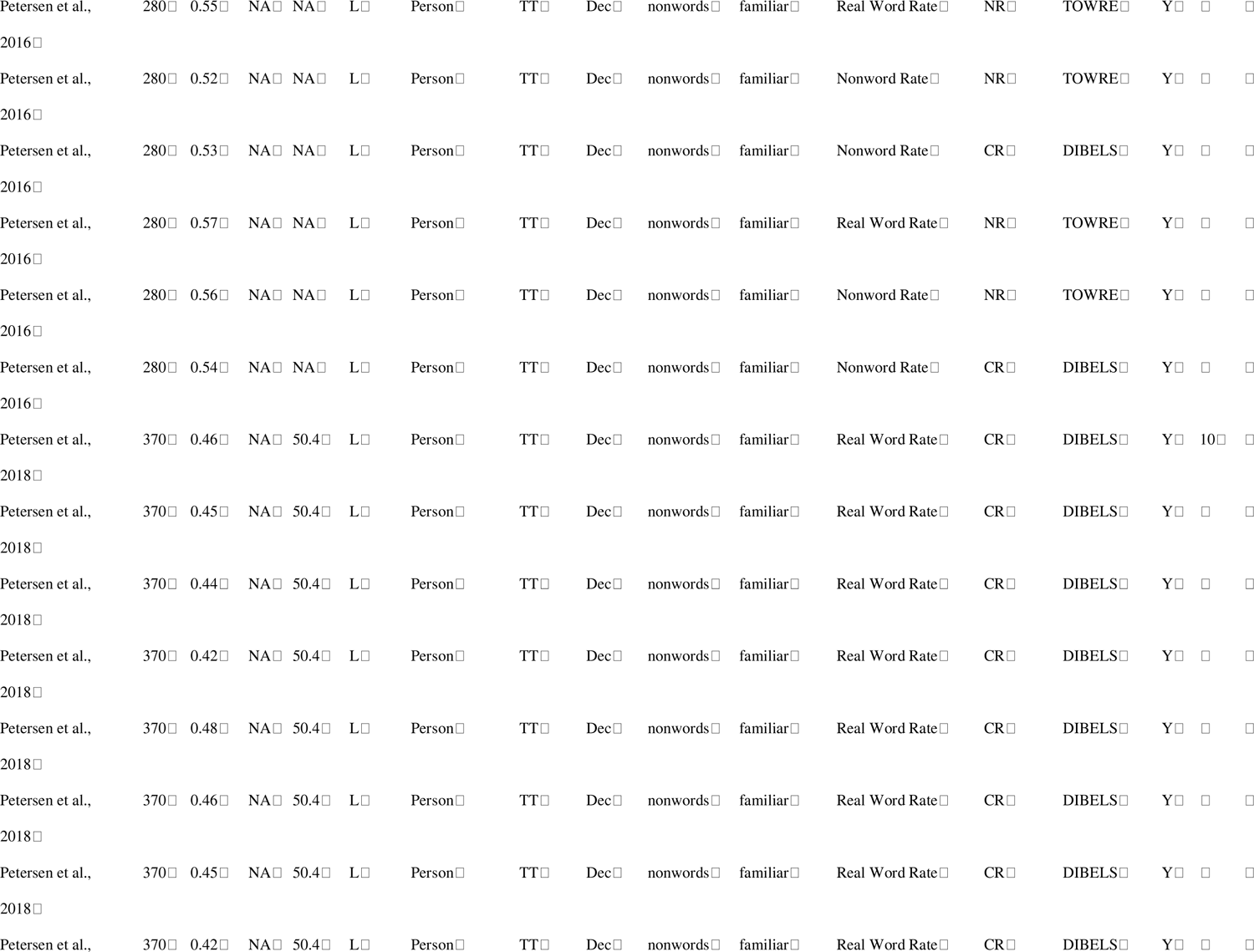
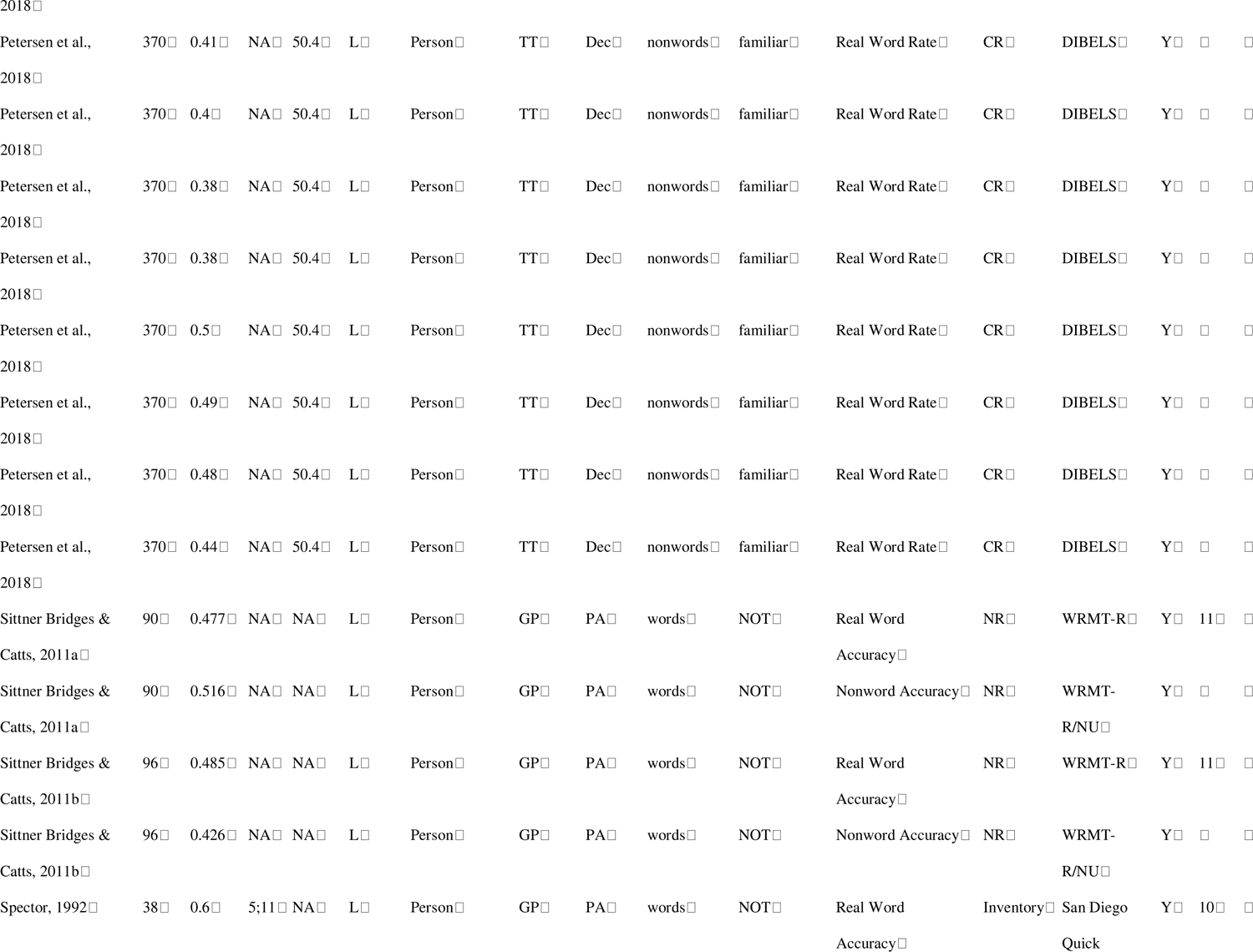
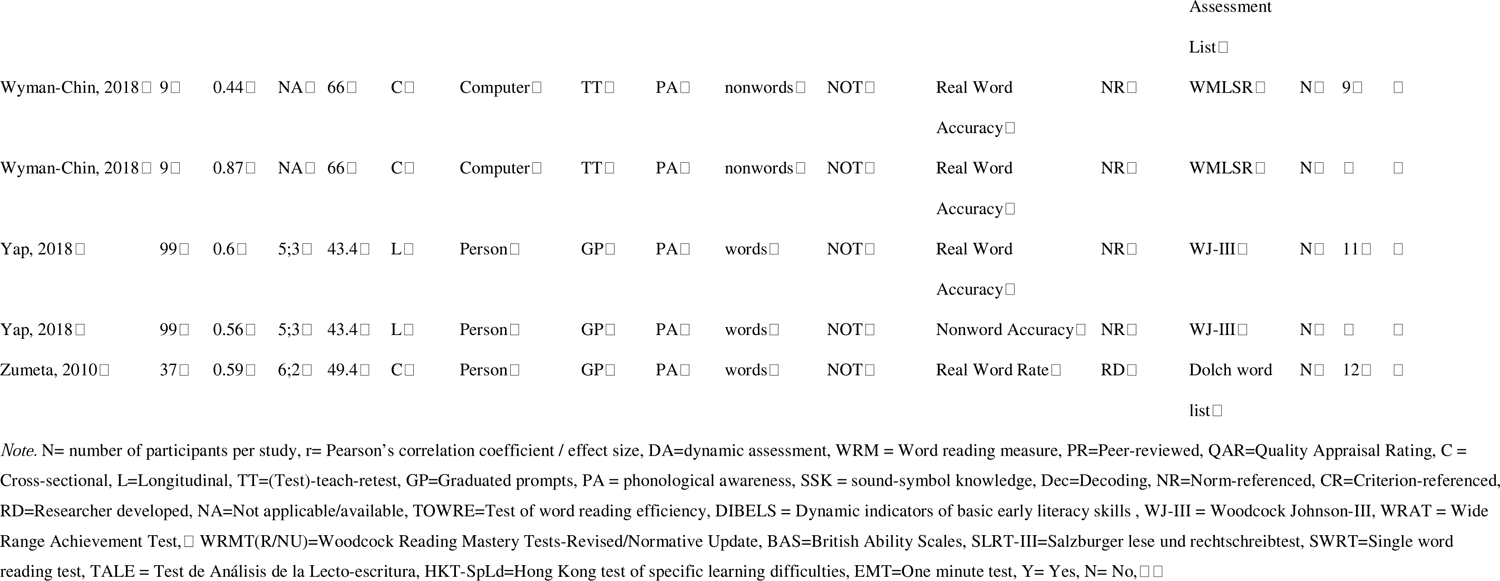
Number of Participants, Effect Size, Mean Age, Grade, % Males, Study Design, Skills Evaluated and Characteristics of DAs and Word Reading Measures of Included Studies.□.

**Table 2.**
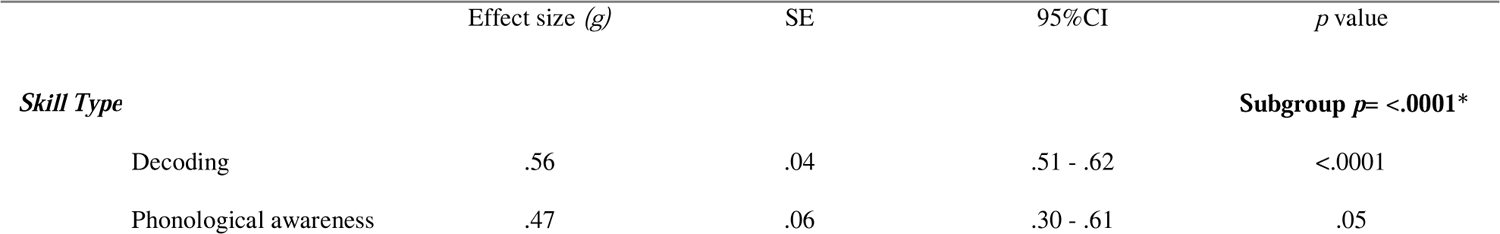
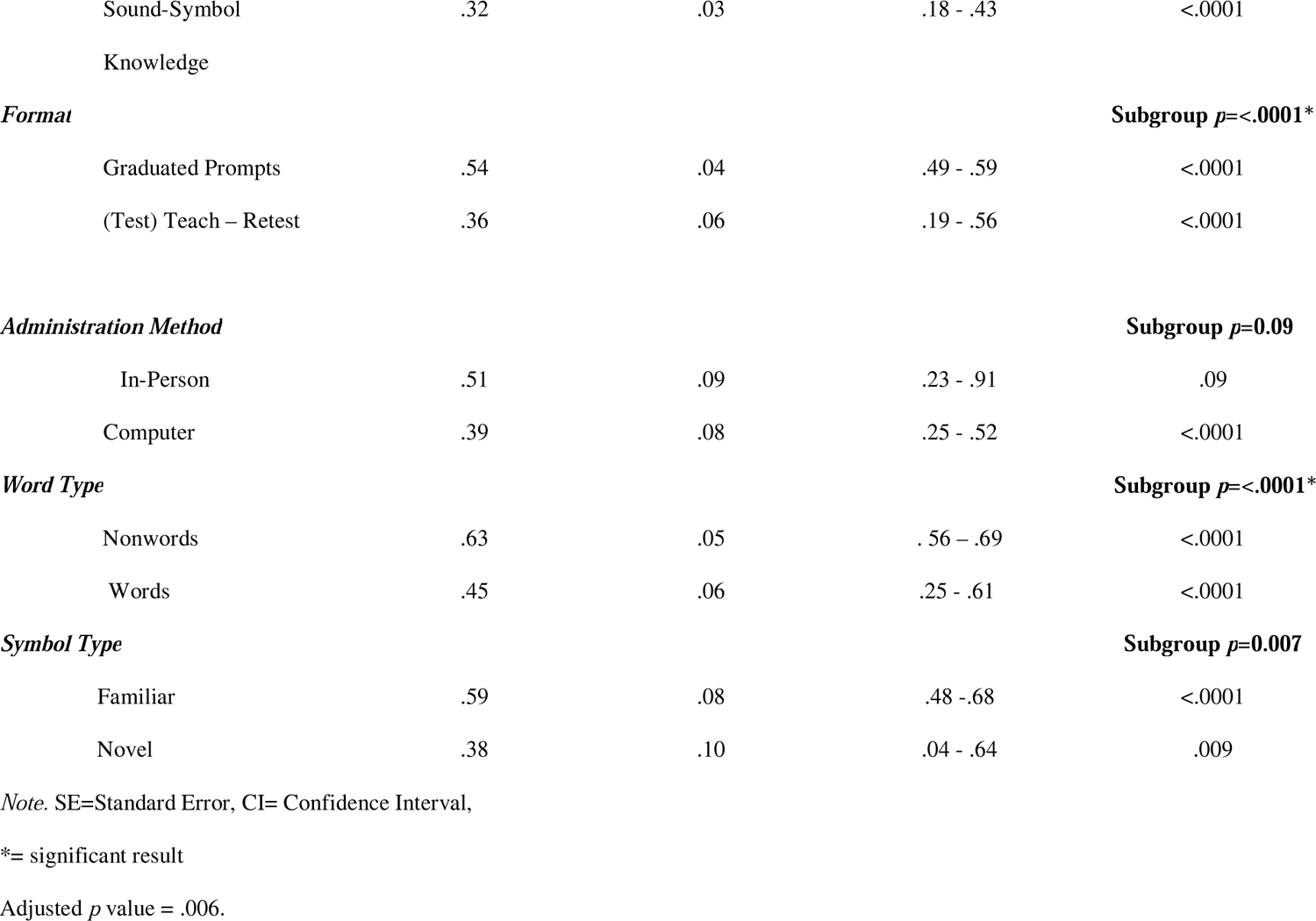
Results of Subgroup Analysis by Skill Type, Format, Administration Method, Word and Symbol Type.

|  | Effect size (*g*) | SE | 95%CI | *p* value |
| --- | --- | --- | --- | --- |
| ***Skill Type*** |  |  |  | **Subgroup $p = <.0001^*$** |
| Decoding | .56 | .04 | .51 - .62 | <.0001 |
| Phonological awareness | .47 | .06 | .30 - .61 | .05 |
| Sound-Symbol | .32 | .03 | .18 - .43 | <.0001 |
| Knowledge |  |  |  |  |
| **Format** |  |  |  | **Subgroup $p=<.0001^*$** |
| Graduated Prompts | .54 | .04 | .49 - .59 | <.0001 |
| (Test) Teach – Retest | .36 | .06 | .19 - .56 | <.0001 |
| **Administration Method** |  |  |  | **Subgroup $p=0.09$** |
| In-Person | .51 | .09 | .23 - .91 | .09 |
| Computer | .39 | .08 | .25 - .52 | <.0001 |
| **Word Type** |  |  |  | **Subgroup $p=<.0001^*$** |
| Nonwords | .63 | .05 | .56 – .69 | <.0001 |
| Words | .45 | .06 | .25 - .61 | <.0001 |
| **Symbol Type** |  |  |  | **Subgroup $p=0.007$** |
| Familiar | .59 | .08 | .48 - .68 | <.0001 |
| Novel | .38 | .10 | .04 - .64 | .009 |

Note. SE=Standard Error, CI= Confidence Interval,
\*= significant result
Adjusted $p$ value = .006.

### Study Selection and Screening Reliability

Study selection and data extraction were managed in Covidence, a web-based software that facilitates completion of reviews (Covidence, 2023). A team of ten research assistants (RAs) assisted in title/abstract screening and full text review. At the title/abstract stage, RAs received a one-hour training session covering key concepts and relevant terms (e.g., defining dynamic assessment and each word reading skill) and subsequently completed 100 practice title/abstract screenings on a mock review prior to screening in earnest. At this stage, two independent team members voted to include or exclude based on whether the title and abstract indicated that the paper evaluated a word reading skill DA. Across all pairs of reviewers, the weighted mean average agreement was 94% and the Cohen’s Kappa coefficient was 0.40 which is characterized as fair (McHugh, 2012). In the full text stage, RAs again received a one-hour training session lead by the first author detailing specific eligibility criteria (e.g., reviewing whether a word reading measure was included, whether the age group was correct, etc.). Each RA completed a practice full text review of a paper with feedback from the first author. Two independent reviewers then voted to include or exclude full texts based on whether they met the pre-defined eligibility criteria. At this stage, interrater agreement across pairs was 85% and the weighted mean Cohen’s Kappa coefficient was 0.66, which is considered substantial (McHugh, 2012).

As demonstrated in Figure 1, 24 articles of the 4824 records identified via the database search were relevant and included. A search of three preprint repositories yielded 850 articles of which one was included. Forward searching of these 25 included articles via Google Scholar led to identification of an additional 9 studies. The reference lists of the 34 articles were reviewed to determine if there were relevant articles that had been missed. One additional study was identified through this process. Finally, callouts were made for unpublished studies or data to mailing lists, via posts to social media and by directly contacting labs conducting literacy related research across Canada, the United State and Europe, but this did not lead to identification of any additional relevant articles. In summary, 35 articles met the criteria for inclusion. The study identification process, including reasons for study exclusion (e.g., no use of a dynamic assessment of one of three word reading skills as in Navarro et al., 2018) is outlined in the PRISMA diagram below (Page et al., 2021).

**Figure 1.**
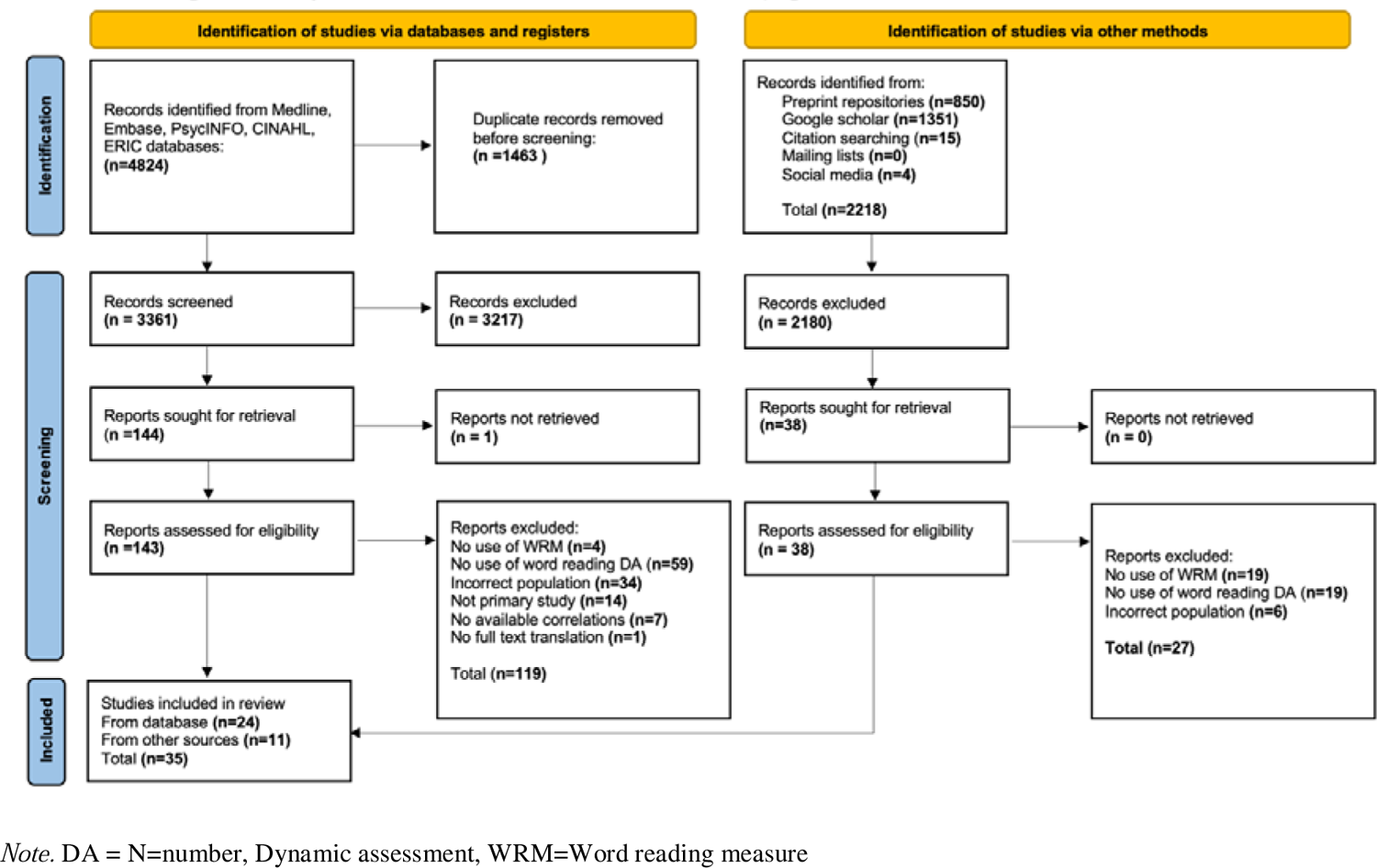
Preferred Reporting Items for Systematic Review and Meta-Analyses Flowchart

### Coding Data Items

Data from relevant articles was extracted using a custom template on Covidence, which is available on the Open Science Framework protocol site (Wood & Molnar, 2022). The first and second author both extracted data from all articles and compared and consolidated findings. Any disagreements were resolved through discussion between all three authors. The following data points were extracted for each included study:

### General Information

The study title, journal name, date of publication, DOI, author name(s), institutional affiliation(s), funding, any potential conflicts of interest, and the country in which the study took place.

### Participant Characteristics

The number of participants included in analyses, the percentage of males, as well as the mean age of the children at the outset of the study was noted. A total of 6683 participants were included. The overall mean age was 5 years 6 months, and the overall average percentage of males was 51%. Mean age was not reported for 14 studies and the percentage of males was not reported for 9. We were able to ensure that studies that did not report mean age still met inclusion criteria, as all minimally reported the grade of participants (e.g., indicated that participants are in grade 1 and therefore between age 4-10). Authors also extracted information regarding participant reading status (typically developing vs. at-risk), language status (monolingual vs. bilingual; and age (4-5, 6-7 vs. 8-9). These factors are examined in a separate paper evaluating the validity of DAs of word reading skills across diverse populations (Wood et al., 2023). Table 1 provides additional details regarding the mean age and % of males of included studies.

### Effect Sizes

Pearson’s correlation coefficients representing the relationship between DAs and WRMs were extracted. All relevant correlation coefficients listed were noted (e.g., a DA that used two PA tasks and their correlations with WRMs). In some instances, a lower score on a DA translated to better performance. In these situations, negative correlations were transformed to positive ones for analysis. From the 35 studies (*k*), a total of 192 effect sizes (*m*) were extracted, with a mean of 5.6 effect sizes per study and a range of 1-30. To account for dependencies between samples, we noted whether two studies conducted analyses on the same set of participants, and if so, nested these effect sizes together under the same study group. For example, Petersen et al., (2018), conducted a follow-up study using the same participant set as in Petersen et al., (2016), and so these effect sizes were grouped together under the umbrella of one study, using robust variance estimation (see statistical analyses below for further details). Study design (e.g., concurrent or longitudinal) was noted. Effect sizes representing the relationship between DAs and WRMs are presented in Table 1.

## Measures

### Dynamic Assessments (DAs)

In this review, DA is defined as an assessment that provides teaching, training, feedback on performance, and/or prompting in testing. In some instances, these measures were not reported as “DAs” but were described as paired associate learning tasks. This was typically for measures that evaluated SSK skills (e.g., Liu & Chung, 2021). We reported the word reading skills evaluated (phonological awareness, sound-symbol knowledge, and/or decoding), and the task used to assess the skill (e.g., phonological awareness/phoneme blending). If multiple tasks were used to evaluate a skill, authors listed all tasks utilized. A DA was characterized as SSK if the task involved learning the relationship between a visual referent (symbol or letter) and a syllable or phoneme. DAs were considered to evaluate PA skills if they assessed one or more of the auditory skills of rhyming, blending, segmenting, manipulating, deleting, substituting phonemes, syllables, words or onset and rimes. Tasks that required a child to recognize more than one symbol-sound relationship and blend these sounds together to “read” multi-symbol words (e.g., CV, VC, CVC etc.) were labelled as decoding tasks. Authors also noted the format of the DA (i.e., graduated prompts (GP) or train/test (TT). DAs were considered GP if a series of prompts was used after a participant response to an individual test item and were characterized as TT if they minimally incorporated a teaching/training phase, followed by a separate static post-test. In terms of administration method (i.e., in person or computer) DAs that were conducted virtually by a clinician over the computer, or those that were computerized (i.e., no clinician) were considered as computer-based administration, while all others were characterized as in-person. For word type, DAs that used words that existed in the language of testing’s lexicon were considered real words (e.g., cat in English) while those that used invented words (e.g., meeb in English), or words from other languages were considered nonwords (e.g., “copa” a Spanish word in an English task). Finally, authors indicated whether novel or familiar symbols were used. DAs that used letters or characters that belonged to the orthography of the language of testing were said to use familiar symbols (e.g., using alphabet letters in an English DA), while those that used invented symbols or letters from orthographies distinct from the language of testing were characterized as using novel symbols (e.g., using Hebrew letters in an English DA measure).

Of the 192 effect sizes from the 35 included studies that examined use of a DA, most evaluated decoding (*k*= 20, *m*=123), some assessed sound-symbol knowledge (*k*=12, *m*=45) and fewest examined phonological awareness (*k*=12, *m*=24). In some studies, the DA evaluated more than one word reading skill, such as in the case of Cho and Compton (2015) where one subtest evaluated SSK and another decoding. In terms of format, 20 studies used a graduated prompts approach (*m*=98), while 15 used a (test)-teach-retest approach (*m*=94). Most DAs were conducted in person, (*k*=24, *m*=156) and fewer via computer (*k*=11, *m*=36). DAs of phonological awareness and decoding tasks used either real or nonwords in their items. Of the studies that evaluated these skills, most used nonwords (*k*=*17*, m=96) while fewer used real words (*k*=15, *m*=51). The studies that evaluated sound-symbol knowledge did not use words given the nature of the task (*k*=12, *m*=45). Decoding and sound-symbol knowledge tasks employed either novel or familiar symbols. Of the studies that evaluated these word reading skills, most used novel symbols (*k*=15, *m*=93) while fewer used familiar symbols (*k*=10, *m*=75). Phonological awareness tasks are auditory in nature and so studies that evaluated this skill did not use symbols (*k*=12, *m*=24). For details about DA characteristics, see Table 1.

### Word Reading Measures (WRMs)

For the purposes of this review, WRMs are assessments that evaluate word reading ability of single real or nonwords using a correct/incorrect grading system and without provision of feedback, prompting or teaching. WRMs were conducted concurrently with the DA or longitudinally at a later timepoint. Of the 35 studies, most effect sizes represented longitudinal relationships (*k*=20, *m*=116) while fewer are associated with concurrent correlations (*k*=, 21, *m*=76). Some studies provided both concurrent and longitudinal correlation coefficients between measures. In extraction, authors noted the name of the WRM, (e.g., the Woodcock Reading Mastery Tests –III), what type of word reading ability was evaluated and whether this was achieved using words or nonwords (e.g., real word accuracy vs. nonword rate). The most used type of WRM was norm-referenced tests (*k*=22, *m*=127), followed by researcher developed tools (*k*=9, *m*=35), criterion-referenced measures (*k*=3, *m*=29) and finally inventories (*k*=1, *m*=1). The Test of Word Reading Efficiency (TOWRE), a norm-referenced test, was the most commonly used tool (*k*=9, *m*=49), along with the Dynamic Indicators of Basic Early Literacy Skills (DIBELS), (*k*=3, *m*=29) and versions of the Woodcock Reading Mastery Tests (WRMT-R/NU), (*k*=10, *m*=28). Other measures like the Woodcock-Johnson-III (WJ-III), were used less frequently (*k*=2, *m*=6). There was variety across task type, with some effect sizes representing correlations between DAs and nonword accuracy measures (*m*=25), real word accuracy measures (*m*=52), nonword rate tasks (*m*=40), real word rate tasks (*m*=49), or a combination or composite of several word reading abilities (*m*=26). Characteristics of WRMs used are detailed in Table 1.

### Quality Appraisal Ratings

Following extraction, included studies were evaluated independently by the first and second author using an adapted version of two quality assessment tools from the Johanna Briggs Institute (Moola et al., 2020). This tool is available on the Open Science Framework protocol page (Wood & Molnar, 2022). Most studies (*k*=26) were peer-reviewed and those that were not (*k*=9) were Masters or Doctoral theses. Studies were assessed on: (i) participant selection, (ii) index assessments (DAs) (iii) reference assessments (WRMs), (iv) flow and timing of the study, (v) statistical analyses. First, coders rated whether the age, sex, and demographic characteristics of the participants were adequately described. The DA domain rating was informed by whether the tool was explained with adequate detail regarding the skills assessed, and the characteristics of the measure. Coders also noted whether the word reading skill(s) employed were developmentally appropriate for the sample population. The reference assessments (WRMs) were also evaluated on their appropriateness of use for the population, and their psychometric properties. To assess flow and timing, coders evaluated whether the analyses included all participants and if not, whether the author(s) provided reasoning for attrition. Lastly, coders considered whether appropriate statistical analyses were conducted.

Overall, the quality appraisal consisted of 8 items to be rated over 5 domains. Items regarding participants, flow and timing, and statistical analyses were assigned one point, while items concerned with the index test (DA) and the reference tests (WRMs) were worth two points due to their greater significance in achieving review objectives. Conflicts were resolved through discussion between all three authors. Quality scores were ranked as either low quality (0-33%), medium quality (34-66%) or high quality (67-100%). Only medium and high-quality studies were included in the analyses. No studies were excluded based on their score. The overall quality appraisal rating for each study is included in Table 1 below. Please refer to Table 3 in the supplemental material for individual ratings for each question for each study.

### Analyses

All statistical analyses were conducted in R using the metafor package (Viechtbauer, 2010; R Core Team, 2021). First, a random effects meta-analysis with robust variance estimation (RVE) was conducted to examine the overall mean effect representing the association between DAs of word reading skills and WRMs. Random effects models assume that variability can stem from multiple sources, both individual sampling error and heterogeneity between studies, This is appropriate in this instance given that studies evaluated populations of different ages, from distinct locations using varied DAs and WRMs (Borenstein et al., 2010). We elected to use RVE because it permits inclusion of multiple effect sizes from a single study, while accounting for dependence between samples (Pustejovsky & Tipton, 2022).

Prior to analysis, the 192 effect sizes were transformed into Z scores using Fisher Z transformation (Corey et al., 1998). Effect sizes are nested within study cluster, with the assumption that they intercorrelate both within and across clusters. A weighted average of these effect sizes was calculated, then transformed back to a Pearson’s correlation coefficient for interpretation as an overall effect (Hedge’s g). We then conducted five subgroup analyses to examine whether the strength of the association between DAs and WRMs differed across the test characteristics of skill type, administration method, format, word and symbol type. These subgroup analyses were planned a priori. To account for multiple comparisons, we used a Bonferroni adjustment to set a new value for significance (*p=*.006). An additional three subgroup analyses were conducted using the same data based on participant characteristics in a separate forthcoming study for a total of 8 subgroup analyses (Wood et al., 2023). The mixed effect model used in subgroup analysis allows us to determine if the mean effect sizes differ significantly based on DA characteristics.

## Results

### Research Question

Do DAs of word reading skills demonstrate consistent validity with word reading measures when stratified by word reading skill type, administration method, format, word, or symbol type? As demonstrated in Table 1, 35 studies reported 192 correlations between a DA of word reading skills (phonological awareness, sound-symbol knowledge, or decoding) and a WRM. The effect sizes from these 35 studies were included in the random effects meta-analysis with robust variance estimation examining the relationship between DAs word reading skills and WRMs. The forest plot of these 192 effect sizes can be found in Figure 1 of the supplemental material. As anticipated, the overall mean effect size is large (*r*=.49, 95%CI = [0.42-0.55]) suggesting that DAs of word reading skills are strongly correlated with WRMs. This result was expected as measures of word reading skills like decoding, phonological awareness and sound symbol knowledge are known to correlate strongly with and be consistent predictors of later word reading ability. The RVE method used in this analysis assumes a consistent correlation (*r*) across study clusters (Pustejovsky & Tipton, 2022). We confirmed that this result was consistent across all values of *r* (.1 - 1.0). Results reported here are for *r=*.8. This three-level model considered variance from sampling (level 1), within study cluster (level 2) and between study cluster (level 3) heterogeneity. The total *I*^2^ was 91.99%. Level 1 sampling error variance was negligible (8.01%), within cluster variance was moderate (30.15%) and between cluster variance was substantial (61.83%), suggesting that the largest proportion of variance is attributed to differences between study clusters. Between study heterogeneity was anticipated given the variation in dynamic assessment characteristics (e.g., administration method, format etc.). This heterogeneity was examined via subgroup analyses.

The contribution of individual effect sizes to heterogeneity was also examined via a Baujat plot, (Figure 2 in the Supplemental Material) (Baujat, 2002). Two effect sizes from Gellert and Elbro (2017a) were identified in the upper right quadrant of the plot, suggesting they contributed significantly to heterogeneity. No plausible reasons for this were identified based on the characteristics of the study. We reran the meta-analysis with these effect sizes removed, but this did not change magnitude of the overall effect (*g*=.49, 95% CI: 0.41 - 0.55, p=<0.0001) or the degree of heterogeneity (*I*^2^=91.99%), and so we elected to retain them in our analysis given that it is not advisable to remove studies without specific reason (Deeks et al., 2022).

### Risk of Publication Bias

A funnel plot was generated to subjectively examine risk of publication bias (See Figure 3 in the Supplemental material). Visual inspection of the funnel plot suggests potential asymmetry. Several studies with small sample sizes and positive findings were identified and included, compared to studies with small sample sizes and negative findings (e.g., Horbach et al., 2018; Loreti, 2015) and are clustered at the bottom right of the funnel. There is a possibility that studies with negative outcomes were not completed, published, or submitted into the grey literature (Lee & Hotopf, 2012). However this may also be simply because the skills evaluated in the DAs included (phonological awareness, sound-symbol knowledge and decoding) are known to correlate with word reading performance, and so negative effects are not anticipated.. Furthermore, despite the apparent visual asymmetry, Egger’s test was calculated and not found to be significant for presence of plot asymmetry (z=1.42, p=0.16) (Egger et al., 1997). Ultimately, there is minimal risk of publication bias in this analysis.

### Subgroup Analyses

Subgroup analyses by DA word reading skill type, format, administration method word and symbol type, were planned a priori and were conducted to examine whether these characteristics have implications for the criterion reference validity of DAs with word reading measures. Mixed effects models were used to examine whether there were significant differences in mean effect sizes for DAs based on these factors. Results of these subgroup analyses are reported in Table 2.

The adjusted significance value was set to *p*=.006 following our Bonferroni adjustment for multiple comparisons. Findings indicate that there are no significant differences in the strength of correlation between DAs of word reading skills and WRMs based on administration method (in-person vs via computer) (*p*=.09) or symbol type (novel vs familiar) (*p*=.007), though the latter approached significance. However, DAs that employed a graduated prompts format were significantly more strongly correlated with WRMs (*g*=.54) than those that used a (test)-teach-retest format (*g*=.36; *p*<.0001). Significant differences were also observed in terms of strength of correlation between DAs and WRMs based on the type of word reading skill used. Though multiple comparisons for each of the three subgroups were not completed, the mean effect sizes for DAs of decoding (*g*=.56) and phonological awareness (*g*=.47) were larger than DAs of sound-symbol knowledge (*g*=.32; *p*<.0001). Finally, outcomes reveal that DAs that use nonwords (*g*=.63) demonstrated significantly larger mean effect sizes than those that used real words (*g*=.45; *p*<.0001).

## Discussion

This review examined whether characteristics of dynamic assessments (DAs) of word reading skills (phonological awareness, sound-symbol knowledge, and decoding) affected the criterion reference validity of DAs of word reading skills as measured by strength of correlational relationship with performance on a word reading measure (WRM). Thirty-five articles met inclusion criteria of evaluating children with a mean age between 4;0 and 10;0 and that reported a Pearson’s correlation coefficient between a DA of a word reading skill and a WRM. As expected, results of the overall meta-analysis suggest that DAs of word reading are strongly correlated with WRMs.

Analysis of *word reading skill type* indicates that performance on DAs of decoding and phonological awareness (PA) is strongly correlated with performance on word reading measures, while performance on DAs of sound-symbol knowledge (SSK) is only moderately correlated. These results are in line with Dixon et al., (2022a), who in their systematic review documented that DAs of PA and decoding generally accounted for greater unique variance beyond static word reading accuracy performance (between 4-21%, and 1-17% respectively), and in nonword reading fluency (between 1-9% and 1-17% respectively), than SSK tasks, which accounted for only 2-6% unique variance in later word reading fluency and accuracy. These findings suggest that in DA, PA and decoding tasks may be better suited than SSK tasks to evaluate current and predict later word reading ability. This contrasts with SA, where letter-sound tasks have been found to demonstrate stronger correlations with word reading performance than PA tasks (e.g., National Early Literacy Panel, 2008). We speculate that the differences in relative strength of correlation between word reading skills of PA and SSK and word reading across static and dynamic domains may be due to the nature of the assessments themselves. In static assessments, children are evaluated on acquired knowledge. PA tasks may be less familiar, and therefore more challenging than SSK tasks. A static SSK task merely requires a child to recognize a letter and name the sound, while a PA task could range in complexity, from something simple like asking the child to segment a word into syllables, to something more challenging like manipulating phonemes in words. Since static PA tasks are often unfamiliar and complex, and children receive no prompting, teaching or feedback in the assessment, many perform poorly on these measures at the earliest stages of learning to read. This results in floor effects, which weaken the predictive value of a measure. Indeed, previous research has documented that static PA tasks demonstrate floor effects for a longer period of time relative to letter naming or letter-sound tasks (Catts et al., 2009). However, in DA, the opposite may be true, given that clinicians and researchers are attempting to not only evaluate what the child already knows, but also how they learn. When presented with an unfamiliar complex task, such as a PA task, there is ample opportunity to evaluate *ability to learn*. However, a simple, more familiar SSK task may not create the opportunities necessary to demonstrate this ability. These results highlight the differences between static and dynamic assessments of early word reading skills. In SA, tasks must be simple enough to ensure that not all children will struggle given their limited acquired knowledge. However, in DA, tasks must be complex enough that children have the opportunity to learn in the context of the assessment. more complex

The analyses evaluating DA *format* found significant differences between the graduated prompts (GP) or train-test (TT) approaches in favour of the GP format. These results are consistent with findings from a previous review which found that DAs that used contingent feedback demonstrated stronger predictive validity than those that used non-contingent feedback (Caffrey et al., 2008). GP DAs are scripted and use contingent feedback, employing a series of pre-defined, increasingly explicit prompts following an examinee’s response (e.g., Spector’s (1992) use of 6 pre-defined prompts). Many of the TT DAs also used non-contingent feedback (e.g., Horbach et al., used non-contingent scripted verbal feedback). However, the feedback in the training and teaching phase of TT DAs was characterized by greater variability. For example, in Petersen and Gillam’s (2015) study examining a DA of nonword decoding, examiners used noncontingent feedback to teach children how to read the nonwords if they were unsuccessful in the initial pre-test phase. This increased variability may have contributed to the weaker relationship between TT DAs and WRMs as the potential for individual differences in teaching styles across examiners and researchers in TT formats is more likely to have had an influence on DA performance.

The analysis examining the role of *administration method* in DAs of word reading skills found that there were no significant differences between those administered in-person, vs. those conducted by a person via computer/computerized. These findings are consistent with a previous review which reported no significant differences between in-person and computer administration methods for static assessments (SAs) in pediatric language assessment (Alfano et al., 2022). Though the difference was not significant, a larger mean effect size was found for DAs administered in person. This could be a result of several factors. First, all WRMs were conducted in-person. It is promising that despite this difference in administration method, computer administered DAs still demonstrated strong mean effect sizes with in-person WRMs. Second, as posited earlier, it is possible that because DAs are characterized by increased interaction between examiner and examinee relative to SAs, administering them via computer may result in a reduced ability to engage in meaningful interaction or provision of accurate feedback. Similar challenges (e.g., technical issues disruption assessment, the need for caregiver support in evaluation and difficulties associated with providing feedback and maintaining child engagement) have been documented in the literature examining virtual use of SAs (e.g., Hodge et al., 2019, Wood et al., 2021). However, results from these studies and this review indicate that much like SAs, computer-based or virtual DAs are a valid alternative to in-person administration.

Regarding *word type* in DAs of word reading skills, results indicate that DAs that use nonwords are associated with significantly greater mean effect sizes than those that use real words. These results differ from previous findings in the SA literature, which suggested that nonwords were too distant from real word reading to be valid and were impractical for beginning readers who lack the necessary skills to decode (Wagner et al., 1997). However, as previously stated, it is possible that use of nonwords in DAs permits evaluation of a child’s ability to learn, since all children are unfamiliar with them, and cannot use previous knowledge or experiences to recognize words in testing (Hoover & Tunmer, 1993). The issue of lacking the necessary skills is also resolved, given that DAs incorporate teaching, feedback and prompting that parallels what a child might receive in a classroom context. This finding again highlights the differences between static and dynamic approaches. In a static approach, familiar material is required to evaluate acquired knowledge, but in DA, unfamiliar material, such as nonwords, may be better suited to evaluate learning potential.

This trend of unfamiliarity leading to increased capacity to evaluate ability to learn in DA was not reflected in the subgroup analysis by *symbol type*. There were no significant differences between DAs that used novel vs. familiar symbols. DAs that used familiar letters or characters were associated with larger mean effect sizes than those that used novel symbols, and this difference approached significance. We speculate that this may be a result of the types of symbols used in these DAs. For instance, some used real letters and characters from a different language in their test items (e.g., Aravena et al., 2013 used Hebrew characters in evaluating Dutch children). Others however, used symbols or that did not resemble any letter or character in an existing script (e.g., Horbach et al., used dots and dashes to represent the syllable-sound correspondences in their DA measure). True letters and characters, whether familiar or unfamiliar, exhibit features and characteristics that allow them to be differentiated from scribbles or symbols (Dehaene, 2009; Heimann, et al., 2013). It is possible that unfamiliar symbols that minimally resemble real characters or letters may be better suited to predict word reading ability.

Despite this, these results still have important implications for the validity of DAs of word reading skills. The use of nonwords and novel symbols has the potential to further reduce bias against children with diverse linguistic experiences. When words and symbols are unfamiliar to all, lack of experience with the oral and print system of the language of evaluation has less impact on performance. Results of this review support the use and development of nonword novel-symbol based DA measures. These types of tools could be used to equitably evaluate children with and without experience in the language of testing. This is in contrast with static tools, which are developed for and can only be used in a valid, unbiased manner with a single language group, typically a monolingual population. A DA of word reading skills that uses nonwords and novel-symbols evaluates ability to learn word reading skills, while minimizing impact of previous linguistic experience. This type of tool and could act as an equitable alternative to language-specific SAs for culturally and linguistically diverse children for whom there are limited assessment tools.

### Limitations

First, while we endeavored to examine relevant DA characteristics and their implications on validity, it is possible that there are other factors that may be contributing to the overall strength of relationship between DAs of word reading skills and word reading measures. One such factor is word reading measure type. In the Caffrey et al., review, (2008), authors examined whether type of outcome measure had implications for validity of DAs and reported that researcher developed tools demonstrated the largest mean effect size relative to norm and criterion-referenced measures, or teacher/clinician ratings. Regrettably, we were not able to replicate the subgroup analysis of word reading measure type fairly, since a large majority (156/192) of effect sizes represented correlations between DAs and a norm-referend or criterion-referenced WRM, while significantly fewer (35/192) used a researcher developed tool, and none used a teacher or clinician rating. Second, correlation coefficients were selected as the measure of effect size because they are consistently reported. While this allowed for inclusion of additional studies, as a result, only correlational inferences can be made about results. Finally, it is possible that relevant studies may not have been identified because they were published in a language that our review team was not able to read (e.g., many studies in Korean and Hebrew were excluded in the title and abstract screening phase), or because they used key terms not captured by our search strategy.

### Clinical Implications

The results of this systematic review and meta-analysis have implications for clinicians like speech-language pathologists and psychologists and educators who routinely evaluate word reading skills. Outcomes suggest that, when possible, clinicians should favour DAs of phonological awareness and decoding skills, that are structured in a graduated prompts format, and that use nonwords comprised of familiar or novel symbols. Results indicate that these measures can be conducted in-person or virtually, which is particularly relevant post-pandemic, as many professionals continue to evaluate children in a virtual context. An example to consider that meets most of these criteria and that is available for free online is the CUBED-3 dynamic decoding measure (DDM). The DDM, a measure developed based on the included studies conducted by Petersen et al., (2016) and Petersen & Gillam (2015), uses a test teach retest approach rather than a graduated prompts approach but evaluates both phonological awareness and decoding, uses nonwords comprised of familiar letters and is administered in person. This measure can be used as a criterion-referenced screening tool, to set intervention targets or to monitor progress (CUBED-3 Dynamic Decoding Measure (DDM), Petersen & Spencer, 2023).

### Tool Development and Suggested Future Research

Results of this study also inform development of novel DAs of word reading skills, or revisions of existing tools. Notably, findings support the virtual administration of DAs of word reading skills. This is an important consideration given that there has been a significant increase in the provision of virtual care and tele-assessment since the COVID-19 pandemic (Campbell & Goldstein, 2021). Researchers should consider developing virtual versions of DAs to ensure that assessment of these critical early reading skills can be made available in any future disruptions to in-person education or clinical services, or simply to ensure that children living in rural or remote areas have access to high quality, equitable dynamic assessments. Study outcomes also support the use of dynamic assessments of word reading skills that use nonwords and novel symbols. Not only does unfamiliarity of words and symbols lend itself well to a DA task where the goal is to evaluate ability to learn, this type of measure may also minimize linguistic and cultural bias associated with traditional static word reading skill tasks. Developing new DAs or revising existing measures to include nonword, novel-symbol based versions is critical for monolingual and bilingual children for whom there are no language specific tests of word reading skills available.

Beyond this, future studies can also directly compare DAs with differing characteristics, using research designs and statistical analyses that permit a better understanding of the causal role these factors play in predicting reading ability and identifying reading disorder at various timepoints in a child’s journey of learning to read. This can be achieved through longitudinal studies comparing the relative predictive validity of DAs that differ in their format, administration method, word and symbol type, or other relevant factors via regression or structural equation modelling. Studies should also explicitly examine whether specific characteristics of DAs of word reading skills have a greater capacity to limit floor effects associated with traditional static measures or result in improved diagnostic accuracy. Ideally, these studies should include populations for whom DA is purported to be most useful, particularly bilingual children and those with limited previous literacy experiences.

## Data Availability

All data produced are available online at https://osf.io/bcghx/

## Acknowledgements

This study was supported by a Canada Graduate Scholarship-Master’s grant from the Social Sciences and Humanities Research Council of Canada, at the Rehabilitation Sciences Institute at the University of Toronto, an Ontario Graduate Scholarship from the Ministry of Colleges and Universities awarded to E. Wood, a University of Toronto Excellence Award awarded to K. Biggs, and a Natural Sciences and Engineering Research Council of Canada grant awarded to Dr. M. Molnar.

