## Supplemental Table 1 for "Characteristics of dynamic assessments of word reading skills and their implications for validity: A systematic review and meta-analysis"

*Search terms for concept 1 – Dynamic assessment*

| Medline | Embase | PsycINFO | CINAHL | ERIC |
| --- | --- | --- | --- | --- |
| No subject heading    (dynamic OR mediat*) ADJ3 (Assess* OR test* OR screen* OR measur* OR tool*).tw,kf.    OR    respon* ADJ3 interven*.tw,kf.    OR    Modifiability ADJ3 Index.tw,kf.    OR    Learning Potential.tw,kf.    OR    (comput* adapt* test*).tw,kf. | *Clinical assessment* OR *Clinical assessment tool* OR *Language test*    dynamic OR mediat*) ADJ3 (Assess* OR test* OR screen* OR measur* OR tool*).tw,kw.    OR    respon* ADJ3 interven*.tw,kw.    OR    Modifiability ADJ3 Index.tw,kw.    OR    Learning Potential.tw,kw.    OR    (comput* adapt* test*).tw,kf. | *Measurement*    dynamic OR mediat*) ADJ3 (Assess* OR test* OR screen* OR measur* OR tool*).tw    OR    respon* ADJ3 interven*.tw    OR    Modifiability ADJ3 Index.tw    OR    Learning Potential.tw    OR    (comput* adapt* test*).tw | *Speech and Language Assessment*    TI (Dynamic N3 (Assess* OR test* OR screen* OR tool* OR task* OR measur* ) ) OR AB (Dynamic N3 (Assess* OR test* OR screen* OR tool* OR task* OR measur* ) )    OR    TI ((Learning potential) N3 (assess* OR screen* OR test* OR tool* OR task* OR measur*)) OR AB ((Learning potential) ADJ3 (assess* OR screen* OR test* OR tool* OR task* OR measur*))      OR    TI response to intervention OR AB response to intervention      OR    TI(comput* Adapt* test*) OR AB(comput* adapt* test*) | *Educational assessment*    (Dynamic) NEAR/3 (assess* OR test* OR screen* OR measur* OR tool*)    OR    (Mediat*) NEAR/3 (assess* OR test* or screen* or measur* OR tool*)    OR    (Respon*) NEAR/3(interven*)    OR    (Modifiability) NEAR/3 (Index)    OR  Learning potential  OR  Comput* adapt * test* |
