## Supplemental Table 2 for "Characteristics of dynamic assessments of word reading skills and their implications for validity: A systematic review and meta-analysis"

*Search terms for concept 2 – Literacy*

| Medline | Embase | PsycINFO | CINAHL | ERIC |
| --- | --- | --- | --- | --- |
| *Literacy* OR *Reading OR* *Writing*    phonem*.tw,kf.    OR    phonolog*.tw,kf.    OR    phonic*.tw,kf.    OR    (sound* ADJ3 (blend* OR segment* OR manipulat* OR substitut* OR delet*)).tw,kf.    OR    ((letter* OR alphabet*) ADJ3 knowledge)tw,kf.    (read OR reading).tw,kf.    OR    (write OR writing).tw,kf.    OR    (spell OR spelling).tw,kf.    OR  (decode OR decoding).tw,kf. | *Literacy* OR *Reading*      phonem*.tw,kw.    OR    phonolog*.tw,kw.    OR    phonic*.tw,kw.    OR    (sound* ADJ3 (blend* OR segment* OR manipulat* OR substitut* OR delet*)).tw,kw.    OR    ((letter* OR alphabet*) ADJ3 knowledge)tw,kw.    (read OR reading).tw,kw.    OR    (write OR writing).tw,kw.    OR    (spell OR spelling).tw,kw.    OR    (decode OR decoding).tw,kw. | *Literacy* OR *Reading* OR *Writing Skills* OR *Academic Writing*      phonem*.tw    OR    phonolog*.tw    OR    phonic*.tw    OR    (sound* ADJ3 (blend* OR segment* OR manipulat* OR substitut* OR delet*)).tw    OR    ((letter* OR alphabet*) ADJ3 knowledge)tw    (read OR reading).tw      OR    (write OR writing).tw    OR    (spell OR spelling).tw    OR    (decode OR decoding).tw | *Literacy* OR *Reading*    TI phonem* OR AB phonem*    OR    TI phonolog* OR AB phonolog*    OR    TI phonic* OR AB phonic*    OR    TI (sound*) N3(blend* OR segment* OR manipulat* OR delet* OR substitut*)    OR    AB sound*) N3(blend* OR segment* OR manipulat* OR delet* OR substitut*)    OR    TI (letter* OR alphabet*) N3 (knowledge OR principle)    OR    AB (letter* OR alphabet*) N3 (knowledge OR principle)    OR    TI ((read OR reading)) OR AB ((read OR reading))    OR    TI ((write OR writing)) OR AB ((write OR writing))    OR    TI ((spell OR spelling)) OR AB ((spell OR spelling))      OR    TI ((decode OR decoding)) OR AB ((decode OR decoding)) | *Emergent literacy* OR *Literacy* OR *Literacy education* OR *Literacy skills* OR *Assessment literacy ...*  Phonem*    OR    Phonolog*    OR    Phonic*    OR    (sound*) NEAR/3 (blend* OR segment* OR manipulat* OR substitut* OR delet*)    OR    (letter*) NEAR/3 (knowledge)    OR    (alphabet*) NEAR/3 (knowledge OR principle)    OR    Write    OR    Writing    OR    Spell    OR    Spelling    OR    Decode    OR decoding |
