## Supplemental Table 3 for "Characteristics of dynamic assessments of word reading skills and their implications for validity: A systematic review and meta-analysis"

| Table 3.  *Quality appraisal of included studies* | | | | | | | | | | |
| --- | --- | --- | --- | --- | --- | --- | --- | --- | --- | --- |
| Study | **Participants** | **Dynamic Assessment** | | **Word Reading Outcome Measures** | | **Flow and Timing** | | **Statistical Analyses** | **Overall Score and /12 (%)** | **Overall Appraisal** |
|  | *Were participant characteristics adequately described?*   *(1)* | *Was the DA adequately described? (2)* | *Was the test appropriate for evaluating word reading skills?*  *(2)* | *Was the WR outcome measure adequately described?  (2)* | *Was the WR outcome measure valid and reliable? (2)* | *Were all participants included in analyses?*  *(1)* | *Were reasons for exclusion or loss to follow up adequately described? (*1) | *Were appropriate statistical analyses used?*   (1) |  |  |
| Aravena et al., 2013 | 1 | 2 | 2 | 2 | 2 | 0 | 1 | 1 | 11(92%) | High |
| Aravena et al., 2018 | 1 | 2 | 1 | 2 | 2 | 1 | 1 | 1 | 11(92%) | High |
| Barker & Saunders, 2020 | 1 | 1 | 2 | 2 | 2 | 0 | 1 | 1 | 10(83%) | High |
| Caffrey, 2006 | 0 | 2 | 2 | 2 | 2 | 0 | 1 | 1 | 10(83%) | High |
| Cho & Compton, 2015 | 1 | 2 | 2 | 2 | 2 | 1 | 1 | 1 | 12(100%) | High |
| Cho et al., 2014 | 0 | 2 | 2 | 2 | 2 | 0 | 1 | 1 | 10(83%) | High |
| Cho et al., 2017 | 1 | 2 | 2 | 2 | 2 | 0 | 1 | 1 | 11(92%) | High |
| Chow, 2014 | 1 | 2 | 2 | 2 | 2 | 2 | 1 | 1 | 12(100%) | High |
| Clayton et al., 2018 | 1 | 2 | 2 | 1 | 2 | 0 | 1 | 1 | 10(83%) | High |
| Compton et al., 2010 | 0 | 2 | 2 | 2 | 2 | 0 | 1 | 1 | 10(83%) | High |
| Coventry et al., 2011 | 1 | 2 | 2 | 2 | 2 | 0 | 0 | 1 | 10(83%) | High |
| Cunningham & Carroll, 2011 | 0 | 2 | 2 | 2 | 2 | 1 | 1 | 1 | 11(92%) | High |
| Edwards, 2020 | 0 | 0 | 2 | 2 | 2 | 1 | 1 | 1 | 9(75%) | High |
| Fuchs et al., 2011 | 0 | 2 | 2 | 2 | 2 | 0 | 1 | 1 | 10(83%) | High |
| Gan et al., 2022 | 0 | 1 | 2 | 2 | 2 | 1 | 1 | 1 | 11(92%) | High |
| Gellert & Elbro, 2017a | 0 | 2 | 2 | 2 | 2 | 0 | 1 | 1 | 10(83%) | High |
| Gellert & Elbro 2017b | 1 | 2 | 2 | 2 | 2 | 0 | 1 | 1 | 11(92) % | High |
| Gellert & Elbro, 2018 | 0 | 2 | 2 | 2 | 2 | 0 | 1 | 1 | 10(83%) | High |
| Gillam et al., 2011 | 0 | 2 | 2 | 2 | 2 | 1 | 1 | 1 | 11(92%) | High |
| Horbach et al., 2018 | 1 | 2 | 2 | 2 | 2 | 0 | 1 | 0 | 10(83%) | High |
| Horbach et al., 2015 | 1 | 2 | 2 | 2 | 2 | 0 | 0 | 1 | 10(83%) | High |
| Law et al., 2018 | 0 | 2 | 2 | 2 | 2 | 1 | 1 | 1 | 11(92%) | High |
| Liu & Chung., 2021 | 1 | 1 | 2 | 2 | 0 | 1 | 1 | 1 | 9(75%) | High |
| Loreti, 2015 | 1 | 0 | 1 | 2 | 2 | 1 | 1 | 1 | 9(75%) | High |
| Osa Fuentes, 2003 | 0 | 2 | 2 | 2 | 0 | 1 | 1 | 1 | 9(75%) | High |
| Petersen & Gillam, 2015 | 1 | 2 | 2 | 2 | 2 | 1 | 1 | 1 | 12 (100%) | High |
| Petersen et al., 2016 | 0 | 2 | 2 | 2 | 2 | 1 | 1 | 1 | 11(92%) | High |
| Petersen et al., 2018 | 0 | 2 | 2 | 2 | 2 | 0 | 1 | 1 | 10(83%) | High |
| Sittner Bridges & Catts, 2011 | 0 | 2 | 2 | 2 | 2 | 1 | 1 | 1 | 11 (92%) | High |
| Spector, 1992 | 0 | 2 | 2 | 2 | 2 | 0 | 1 | 1 | 10(83%) | High |
| Wyman Chin, 2018 | 0 | 1 | 1 | 2 | 2 | 1 | 1 | 1 | 9(75%) | High |
| Yap, 2018 | 1 | 2 | 2 | 2 | 2 | 0 | 1 | 1 | 11(92%) | High |
| Zumeta, 2010 | 1 | 2 | 2 | 2 | 2 | 1 | 1 | 1 | 12(100%) | High |
| *Note.* DA= Dynamic assessment, WR = word reading, Low = 0-33%, Medium=34-66%, High=67-100% | | | | | | | | | | |
