## Supplementary figures and images for "Characteristics of dynamic assessments of word reading skills and their implications for validity: A systematic review and meta-analysis"

### Supplemental Figure 1

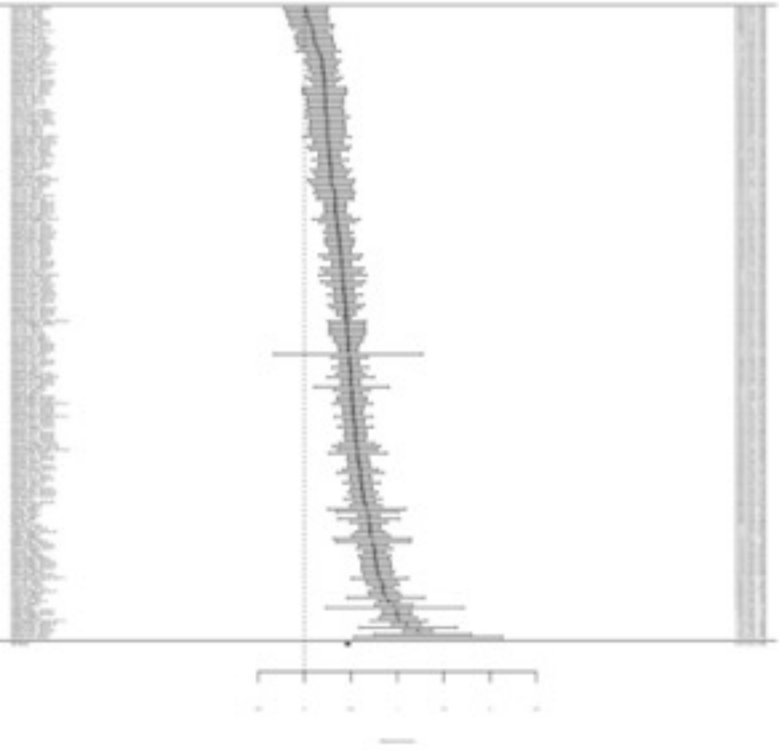

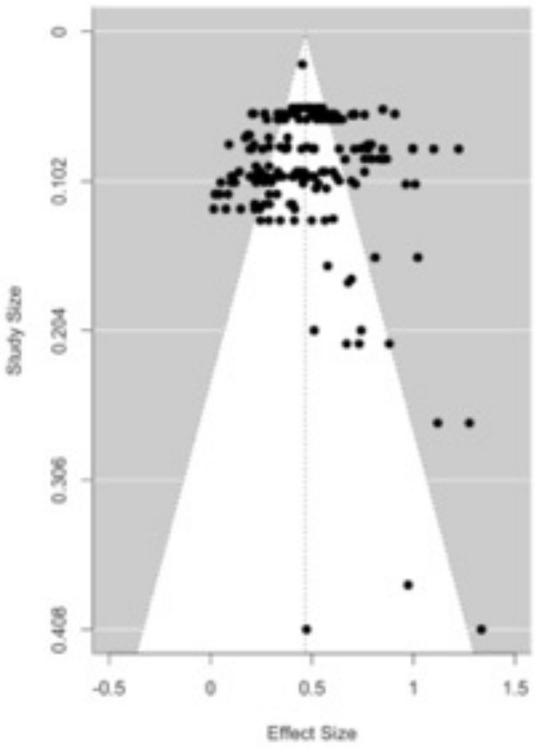

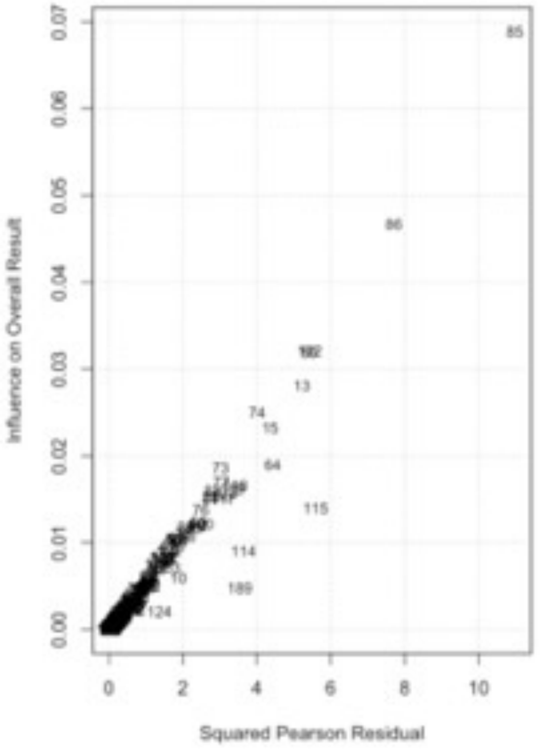

### Supplemental Figure 2

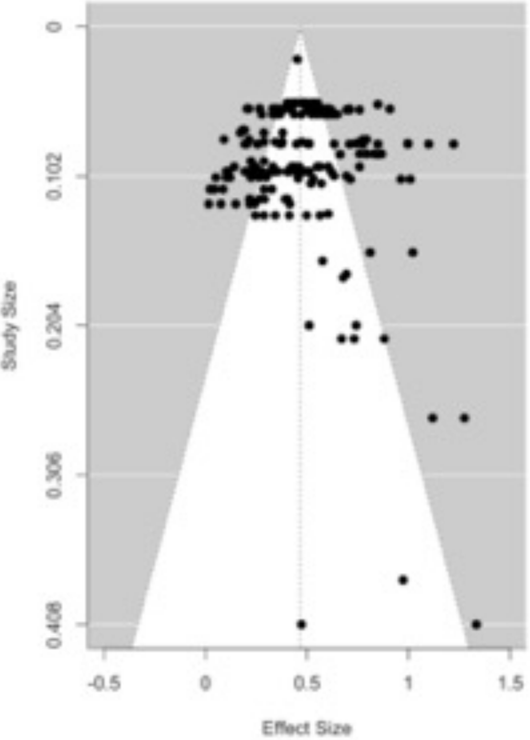

### Supplemental Figure 3

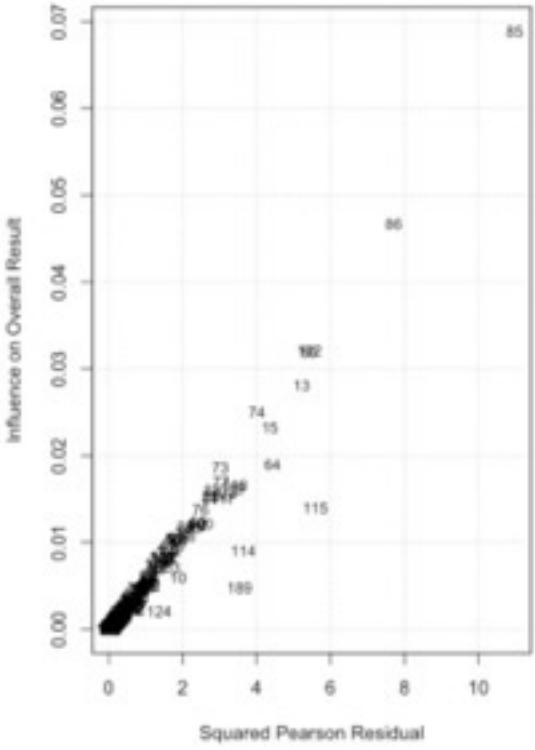
